# Large-scale GWAS meta-analysis of serum antibody levels in healthy individuals reveals distinct genetic architectures

**DOI:** 10.1101/2025.10.25.25338738

**Authors:** Thomas W. Willis, Effrossyni Gkrania-Klotsas, Nicholas J. Wareham, Eoin F. McKinney, Paul A. Lyons, Kenneth G.C. Smith, Chris Wallace

## Abstract

Antibodies are the principal effector proteins of humoral immunity. Dysregulated antibody production is a feature of a number of heritable immune-mediated diseases, such as the antibody deficiencies and IgA nephropathy. To characterise the common-variant architecture of serum antibody levels in the general population, we conducted the largest GWAS meta-analyses to date of serum IgA, IgM, and IgG, attaining sample sizes of 85,204, 55,368, and 58,777, respectively. We identified 82 novel associations across three isotypes, including 38 novel IgA associations (of a total of 77), 36 novel IgM associations (of a total of 82), and 8 novel IgG associations (of a total of 14). We found that isotype-specific genetic architecture was largely disjoint with few signals colocalising across isotypes in spite of their overall positive genetic correlation. We identified a large number of colocalisations between antibody phenotypes and immune-mediated diseases, and 16 between antibody phenotypes and a lymphocyte count phenotype. We report a greatly expanded catalogue of serum antibody-associated variants and characterise these in terms of their pathway context and relation to immune pathology.

## Introduction

Antibodies are soluble immunoglobulins which form the basis of humoral immunity. They are antigen-binding effector proteins responsible for the neutralisation of pathogens and toxins, opsonisation, and the activation of complement. Immunoglobulins exist also as the membrane-bound antigen-binding moiety of cell-surface B-cell receptors (BCRs). In both guises, immunoglobulins are distinguished by the great diversity of antigen-binding affinities they provide to the adaptive immune system, together with T-cell receptors. This diversity depends upon extensive germline polygeny and polymorphism at the *IGH*, *IGK*, and *IGL* loci. This germline-encoded variation is supplemented by somatic hypermutation in the germinal centre.

In addition to its contribution to antibody repertoire diversity, germline genetic variation also contributes to interindividual variation in the quantity of serum antibody produced (1). Heritable influences on antibody production have long been studied in the setting of disorders of antibody production: these include antibody deficiencies, such as Bruton’s agammaglobulinaemia (2) and common variable immunodeficiency (3), and hypergammaglobulinaemias, in which there is an excess of antibody production, such as the ‘hyper IgM’ syndromes (4). Family-based studies in healthy individuals have produced evidence of a substantial heritable component to antibody production (1,5) and the advent of genome-wide association studies (GWAS) has allowed discovery of common variants underlying this heritability (6–9).

Antibody dysfunction and the dysregulation of antibody production feature prominently in the pathogenesis of certain diseases such as IgA nephropathy, IgA vasculitis, and the aforementioned antibody deficiencies. Study of the common-variant architecture of antibody production in healthy individuals can inform understanding of the genetic basis of these diseases (6,10) as well as variation in the response to vaccination in the population at large. In this work we investigate this genetic architecture with two new GWAS of serum IgG and IgM, and large-scale meta-analyses of IgG, IgM, and IgA. We greatly increase the number of common-variant associations, and report updated estimates of serum antibody level heritability, and the genetic correlation of isotypes with one another and immune-related traits. We find, contrary to our expectation and to positive phenotypic and genetic correlations overall, that the majority of significant signals detected in these large meta-GWAS are isotype-specific.

## Material and methods

### GWAS of serum IgG and IgM in the EPIC Cohort

We performed GWAS of serum IgG and IgM in 7,946 and 8,008 individuals, respectively. Participants were drawn from the European Prospective Investigation into Cancer (EPIC)-Norfolk Study (https://doi.org/10.22025/2019.10.105.00004). The study was carried out according to the principles of the Declaration of Helsinki, participants provided informed consent, and the study was approved by the Norwich Local Research Ethics Committee. The GWAS were performed using the methodology described previously in our study of serum IgA in EPIC (10). Briefly, IgG and IgM were quantified using immunosorbent assay; IgG was measured as the sum of the IgG1, IgG2, IgG3, and IgG4 subclasses. Serum antibody measurements were log-transformed and mean-centered. GWAS was performed for each isotype using an additive model in SNPTEST (v2.5.4-beta3) incorporating age, sex, and scores on the first ten genetic principal components.

### Acquisition of public GWAS data sets

We downloaded immunoglobulin GWAS summary statistics (6,11–14) from the EBI GWAS Catalog where available or from URLs provided in individual publications (Supplementary Table 1). Summary statistics from the Eldjarn study were available both with and without median normalisation; we chose to use median-normalised datasets to match the median normalisation of the Pietzner datasets. Liu et al. published two versions of their IgA meta-analysis distinguished by the inclusion or exclusion of data from a study of serum IgA in the deCODE cohort (15). To prevent the inclusion of samples overlapping with those in the Eldjarn study in our own meta-analysis, we used the Liu meta-analysis excluding the deCODE data.

GWAS summary statistics for immune-related traits and diseases (7,16–27) were downloaded from the URLs in Supplementary Table 2.

### Harmonisation and standardisation of GWAS data sets

Prior to downstream analysis, we processed those GWAS datasets which were not available in a harmonised format from the GWAS Catalog with version 1.1.10 of the EMBL-EBI GWAS summary statistics harmoniser (28). This process lifted over variant coordinates to the GRCh38 assembly, inferred the strand orientation of palindromic variants, and aligned alleles to the Ensembl 95 reference.

The phenotypic values used to produce each GWAS dataset were subject to transformation and standardisation by their respective authors, but the method chosen differed between studies (Supplementary Tables 1 and 3). These differences produced phenotypic values, and therefore effect estimates and standard errors, on different scales. To correct for this we estimated the standard deviation of the phenotype in each dataset, exploiting the following relationship between phenotypic variance *var(Y)*, genotypic variance *var(X)*, the sample size *n*, and the variance of the effect estimate *var(*β):

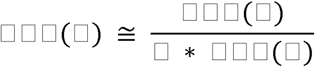

where genotypic variance is given by:

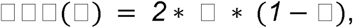

with *f* the minor allele frequency estimated using 1kGP European samples. We regressed *n * var(X)* on the reciprocal of *var(*β) and obtained an estimate of *var(Y)* as the regression slope. We conducted 10 replicates of this regression in each dataset using 10,000 SNPs randomly selected after LD pruning using *PLINK* version 2.00a5.10 (29,30) with a 1,000 kb window, a step size of one, and r^2^ < 0.2. The estimate thus obtained was stable across replicates (Supplementary Table 3). We used the square root of the median of the estimates thus obtained as the estimated standard deviation, which we used in turn as a scaling factor to divide effect estimates and standard errors in each dataset prior to meta-analysis. Our application of this method of standardising summary statistics assumes homogeneity of the underlying phenotype variance across component GWAS; in the absence of phenotype data from these studies, it was not possible to evaluate this assumption.

We did not apply a genomic control correction nor an LDSC intercept correction to the standard errors of the input GWAS nor our final meta-analyses (31).

### GWAS meta-analysis

We performed an outer join between the SNPs of each GWAS for each antibody isotype and then a fixed-effect, inverse variance-weighted meta-analysis.

We designated as candidate ‘lead SNPs’ those variants which had a p-value < 5×10^-8^ and the smallest p-value in the 1Mb interval centered upon the variant. We visually inspected the surrounding 4Mb interval using *locuszoomr* (32) and excluded lead SNPs which appeared to be in LD with a more significantly associated SNP. Estimates of r^2^ reported between pairs of variants in the text were obtained from 1kGP European samples using the LDlink web application (33).

In order to identify novel associations, we downloaded known associations reported in the EBI GWAS Catalog on February 4th 2025 (^28^). We added associations from the GWAS performed by Gudjonsson et al. (34), which we omitted from our meta-analyses due to the possibility of sample overlap with the Icelandic deCODE cohort studied by Eldjarn et al. For IgA, we added associations reported in Table 2 of the study by Liu et al. after lifting them over to GRCh38 and the associations we reported in our earlier meta-analysis of IgA (10). Lastly, we added associations present in the studies included in our meta-analyses, excepting the studies of IgG and IgM in the EPIC Cohort which we publish here for the first time. For each of our isotype-specific meta-analyses, we then merged the list of lead SNPs from signals attaining genome-wide significance with our compiled list of known associations using an ‘overlap join’: where no existing association’s reported lead SNP lay within 500kb window up- or downstream of the lead SNP in our own meta-analysis, we designated the latter as a novel association.

We downloaded the 2024 edition of the IUIS’ classification table of inborn errors of immunity (35) from the IUIS website (https://iuis.org/committees/iei/) and obtained the HGNC symbol and coordinates of the genes listed in this table from Ensembl using biomaRt (36,37). We mapped SNPs to IEI genes which lay within 200kb.

### Estimation of trait heritability and the genetic correlation

We used the *SumHer* component of *LDAK* version 5.1 to estimate heritability and the genetic correlation between traits (38). We used the ‘Human Default’ heritability model when estimating the trait heritability, and the ‘LDAK-Thin’ heritability model and ‘SumHer-GC’ model of confounding when estimating the genetic correlation.

We identified highly significant and possibly spurious association signals in the *IGH* and *IGK* loci (Results). We removed these loci (chromosome 14, positions 105,586,437 to 106,879,844, and chromosome 2, positions 88,857,361 to 90,235,368 in the GRCh38.p14 assembly, GenBank Gene IDs 3492 and 50802, respectively) and the *IGL* locus (chromosome 22, positions 22,026,076 to 22,922,913, GenBank Gene ID 3535) for the purposes of heritability and genetic correlation estimation. For the *IGH* and *IGK* loci, it was necessary to exclude a larger region, 103,000,000 to 107,00,000 and 88,700,000 to 90,235,368, respectively to fully ablate the association signals. We report heritability and genetic correlation estimates with and without inclusion of these loci.

We also report heritability estimates with and without inclusion of the major histocompatibility locus (MHC), which we defined liberally as the interval on chromosome 6 between positions 24,000,000 and 45,000,000. The MHC was excluded from datasets for the purpose of genetic correlation estimation.

We estimated the phenotypic correlation of isotype-specific serum antibody levels using data from the EPIC Cohort. We used Pearson’s correlation for isotype pairs using both raw and log-transformed data, and calculated bootstrap confidence intervals for each estimate using 1,000 replicates.

We used SNP genotype data from 535 European-ancestry samples from the Phase 3 release of the 1000 Genomes Project (1kGP) (39) for out-of-sample estimation of the squared correlation of SNP genotypes (*i.e.* r^2^) and the calculation of *SumHer* SNP taggings. All but one GWAS used in our analyses were performed in European-ancestry cohorts (Supplementary Table 1). The exception, the serum IgA meta-analysis of Liu et al. which combined 16 ancestry-defined cohorts spanning European, African, East Asian, and admixed ancestries, nonetheless consisted largely of European-ancestry individuals (35,094 of 41,263, ∼85%). We therefore identified 1kGP European-ancestry samples as the most suitable reference panel.

We downloaded data from the high-coverage GRCh38 1kGP release (40) and processed it using *PLINK* version 2.00a5.10 to meet the following criteria: SNPs only; MAF > 0.005; missingness per SNP < 0.01; missingness per sample < 0.01; exact test of Hardy-Weinberg equilibrium has p-value > 1e-50. We enforced alignment of reference and alternative alleles to the GRCh38 reference sequence.

### Annotation of GWAS lead SNPs

We assigned possible causal genes to each lead SNP using the Open Targets Platform (41) and Ensembl version 113 (42). Genes were assigned to a variant if the variant was missense in the gene or if the variant was in the fine-mapping credible set for any molecular QTL for the gene. For lead variants with no gene thus assigned, we assigned the nearest gene identified using Ensembl annotations. We then performed pathway analysis to identify biological pathways related to these genes as a whole, and colocalisation between the different antibody phenotypes, with a lymphocyte count phenotype, and with a set of 13 immune-mediated diseases, as detailed below.

### Pathway analysis

To identify biological pathways highlighted by our genetic associations, we first created a test set of genes from the union of possible causal genes across our meta-analyses of IgA, IgG and IgM. We then used the *enrichKegg* function from version 4.14 of the *clusterProfiler* package (43), and the *enrichPathway* function from version 1.50.0 of the *reactomePA* package (44) to test for association with pathways annotated by KEGG and Reactome, respectively.

### Colocalisation

We carried out colocalisation analysis using the *coloc::coloc.abf* function in *coloc* version 5.2.4 (45). In accordance with package defaults we defined the prior probability of a random SNP’s being causal for one trait in a pair but not the other as 1×10^-4^ and the prior probability of a random SNP’s being jointly causal for both traits as 1×10^-5^. We identified candidate serum antibody loci for colocalisation by searching for pairs of lead SNPs for distinct isotypes which lay within 1Mb of one another. For each pair of lead SNPs thus identified, we included in the analysis all SNPs lying in the interval between the lead SNPs as well as those in the 250kb flank regions lying up- and downstream of the same interval. For colocalisation of loci for serum antibody and non-serum antibody traits, we searched a 1Mb interval centered on each serum antibody lead SNP for SNPs reaching genome-wide significance (p < 5×10^-8^) for the non-antibody trait. Where such SNPs were present, we carried out colocalisation using the SNPs in the 1Mb interval. We examined regions where the posterior probability of a shared causal variant exceeded 0.5 using the *coloc::plot_extended_dataset* function, and designated colocalisations on the basis of the posterior probability of a shared causal variant, the posterior odds of the same hypothesis, and the visible concordance of the association signal in each GWAS. The smallest posterior probability of colocalisation for a locus thus identified as the site of a colocalisation was 0.69.

Our serum antibody meta-analyses incorporated multiple studies which differ in the panel of SNPs for which they reported summary statistics. As a consequence, the sample size of each meta-analysis varied across SNPs. Approximate Bayes factors do not account for sample size imbalance of this kind (46) and the resulting miscalibration of fine-mapping (47) may propagate to colocalisation analysis. We conducted a sensitivity analysis, filtering the SNPs included in colocalisation analysis by their sample fraction, *i.e.* the SNP-specific sample size expressed as a proportion of the largest sample size in the meta-analysis. We retained only those SNPs for which the sample fraction in each meta-analysis differed by no more than 0.1. We found that filtering rarely affected the reported posterior probability of a shared causal variant, with two exceptions (Supplementary Figures 1); one of these exceptions, the association signal for IgG and IgA at *TNFSF13*, saw the posterior probability of a shared causal variant differ by ∼1 between the filtered and unfiltered datasets (Supplementary Figures 2 and 3). We disregarded this locus from further consideration as the site of a colocalisation.

## Results

We conducted fixed-effect, inverse variance-weighted meta-analyses of IgG, IgM, and IgA datasets (Figure 1, Supplementary Table 1), attaining sample sizes of 55,368, 58,777, and 85,204, for these isotypes, respectively.

**Figure 1.**
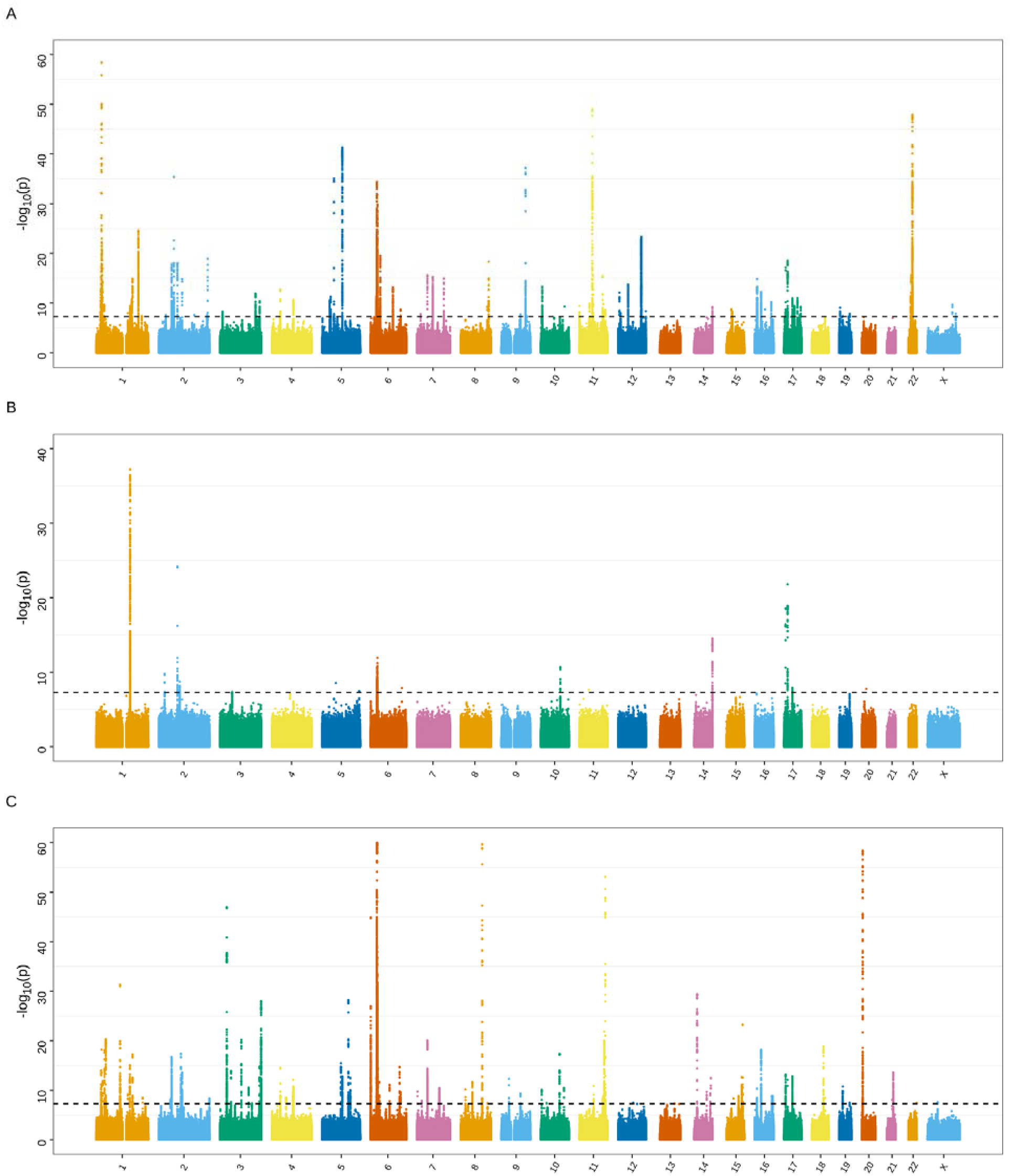
Manhattan plots of GWAS p-values for the meta-analyses of serum IgA (A), IgG (B), and IgM (C). The serum IgA meta-analysis comprised 85,204 samples, the IgG meta-analysis 58,777, and the IgM meta-analysis 55,368. Association signals in the *IGH* region have been omitted for the reason of the large scale of their p-values relative to those in other association signals.

Both the antibody-based EPIC study and aptamer-based Pietzner and Eldjarn GWAS of serum IgA and IgG featured genome-wide significant association signals centered upon the constant region of the *IGH* locus. The serum IgA GWAS replicated the known association of the *IGHA1* missense variant, rs117775520, with serum IgA (15), although the direction of effect differed between the EPIC GWAS and the aptamer-based GWAS (Supplementary Figure 4 and Supplementary Table 4). Variants in the locus were associated with serum IgG in the EPIC, Pietzner, and Eldjarn GWAS but the location of the lead SNP differed substantially between EPIC and the other datasets (Supplementary Figure 5). The Eldjarn GWAS of serum IgM identified a significant but more modest association with rs10873559, near *IGHA1* (Supplementary Figure 6). rs232230, a missense variant in *IGKC*, was associated with serum IgG in the Eldjarn study (Supplementary Figure 7). In our meta-analyses, all three isotypes had associations with variants in the *IGH* locus (Supplementary Figure 8) whereas only IgG and IgA had significant associations in the *IGK* locus (Supplementary Figure 9).

We could not definitively account for the differing locations and effect directions estimated at *IGH* variants in the serum IgA and IgG GWAS. We hypothesised that coding variation within them (e.g. rs117775520) could modify the immunoglobulin epitopes bound by the capture antibodies and aptamers used in affinity-based quantification, and thus induce spurious genotype-dependent variation in antibody measurement. The immunoglobulin allotypes defined by certain coding variants were originally characterised in terms of their differing reactivity to immunologic reagents (48) and recent work has shown IgG1 allotypes do indeed affect detection by commercially available anti-IgG1 antibodies (49). We therefore took a conservative approach, and removed the *IGH*, *IGK* and *IGL* regions from some primary analyses that follow. We report heritability and genetic correlation estimates with the inclusion of these regions for sensitivity purposes.

Outside of the *IGH* and *IGK* loci, we identified 14 associations with serum IgG, of which 8 were novel (Supplementary Table 5), 82 associations with serum IgM (36 novel, Supplementary Table 6) and 77 associations with serum IgA (38 novel, Supplementary Table 7), making for a total of 82 novel associations across all three isotypes (Table 1). We obtained heritability estimates for each isotype using these data (Supplementary Table 8), finding larger estimates for IgA (h^2^ = 0.16) and IgM (h^2^ = 0.21) than for IgG (h^2^ = 0.07). Inclusion of the *IGH*, *IGK*, and *IGL* regions increased serum IgG heritability markedly (21.8%), and IgA and IgM heritability modestly (1.5% and 1.6%). We also examined the influence of the major histocompatibility complex (MHC) upon heritability estimates. The MHC features long-range LD patterns and typically evinces strong genetic associations with immune-related traits; SNPs in the MHC featured associations with all three isotypes in our meta-analyses (Figure 1). Inclusion of the MHC increased estimated IgM heritability the most (22.7%), and had an appreciable effect on IgA (11.0%) and IgG (16.6%), also.

**Table 1.** Lead SNPs from novel genome-wide significant associations in the IgA, IgG, and IgM meta-analyses. ‘MAF’ is the minor allele frequency computed from data in the component GWAS where this information was provided in the original studies. ’Gene(s)’ gives the gene(s) with the most evidence linking it/them to the association signal; the procedure by which we assigned genes to variants, and identified quantitative trait loci (QTL) and missense variants is described in Methods. ‘Missense gene’ reports the gene (if any) in which the lead SNP was identified to be a missense variant. ‘Q’ is Cochran’s Q test statistic for which the degrees of freedom are also given. ‘I2’ is the I^2^ statistic.

All serum immunoglobulin measures were positively correlated on the log scale (Supplementary Table 9). Concordant with this finding were significant positive genetic correlation (r_g_) estimates between serum IgA and IgG, and IgG and IgM, when the *IGH*, *IGK*, and *IGL* loci were excluded (Table 2 and Supplementary Table 10). These r_g_ estimates were reduced in magnitude and made non-significant with inclusion of the *IGH*, *IGK*, *IGL* loci. We attribute this to the potential false positive results in the *IGH* locus sufficing to dilute evidence of pleiotropy in the rest of the genome as measured by the genetic correlation.

**Table 2.** Estimates of the genetic correlation between serum antibody isotype levels. p-values were calculated from a chi-squared test of non-zero genetic correlation. The MHC and *IGH*, *IGK*, and *IGL* loci were excluded from the datasets used to produce these estimates.

| First isotype | Second isotype | Genetic correlation estimate | Standard error | p-value |
| --- | --- | --- | --- | --- |
| IgA | IgM | -0.03 | 0.03 | 3.51E-01 |
| IgA | IgG | 0.21 | 0.04 | 6.10E-07 |
| IgG | IgM | 0.15 | 0.05 | 1.18E-03 |

In spite of the genetic correlation we observed when omitting these loci, however, we found only one locus at which there was strong evidence of a causal variant shared between IgG and IgA (Supplementary Figure 10; posterior probability of colocalisation, ‘PPC’, 0.85), and none at which this was true for IgG and IgM (Supplementary Table 11). This may be explained in part by the much smaller number of IgG associations, consistent with its relatively lower heritability estimate (Supplementary Table 8).

In contrast, although IgA and IgM did not show significant genetic correlation overall, six associations with each isotype showed evidence of a causal variant common to both isotypes in colocalisation analysis (Supplementary Table 11, Supplementary Figures 11-16, minimum PPC = 0.80). For five of these colocalising associations, the direction of effect on serum IgA and IgM was concordant. The exception was on chromosome 4 near *RHOH,* for which association the IgA and IgM lead SNPs had discordant effects on the level of each isotype. *RHOH* encodes the haematopoiesis-specific RhoH GTP-binding protein (50) and mutations in the gene cause an autosomal recessive combined immunodeficiency (51). RhoH-deficient mice show normal IgM production but impaired production of class-switched IgG1 in response to immunisation with T cell-dependent antigen (52), suggesting the *RHOH*-mapped variants may affect serum antibody levels through an impact on the efficiency of class switch recombination. A combination of shared signals, some with the same effect direction (immunoglobulin production) and others with the opposite (class switching efficiency), could explain the lack of overall genetic correlation between these isotypes.

The colocalising associations with concordant direction of effect included a novel association with IgA (lead SNP rs2872516) which colocalised with a known association of the same locus with IgM (rs4795397, PPC = 0.95). However, another novel association with IgG (rs17676923) in this region appeared to relate to a distinct causal variant with a posterior probability of distinct causal variants of ∼1. This was supported by an examination of LD in the region which showed that the lead SNPs for IgA and IgG were not in LD (r^2^ = 0.03), although those for IgA and IgM were (r^2^ = 0.83). All three SNPs belong to eQTL credible sets for *GSDMA*, *GSDMB*, and *IKZF3* in blood.

Given the overlap of association signals, we used pathway analysis to summarise the associations seen across all isotypes. This identified six and ten significant KEGG and Reactome pathways, respectively (q-value < 0.05, Figure 2), the most prominent amongst them relating to B cell receptor and NF-kB signalling. Our gene selection strategy was deliberately broad, which sometimes leads to the nomination of multiple genes for a single association. By comparing this list of genes with the list of genes in associated pathways we can highlight more likely causal genes in some cases (Supplementary Figure 17). Of note, we see that the associated pathways typically contain associations with more than one isotype, highlighting the convergent biology of the GWAS associations in spite of the limited evidence of shared signals.

**Figure 2.**
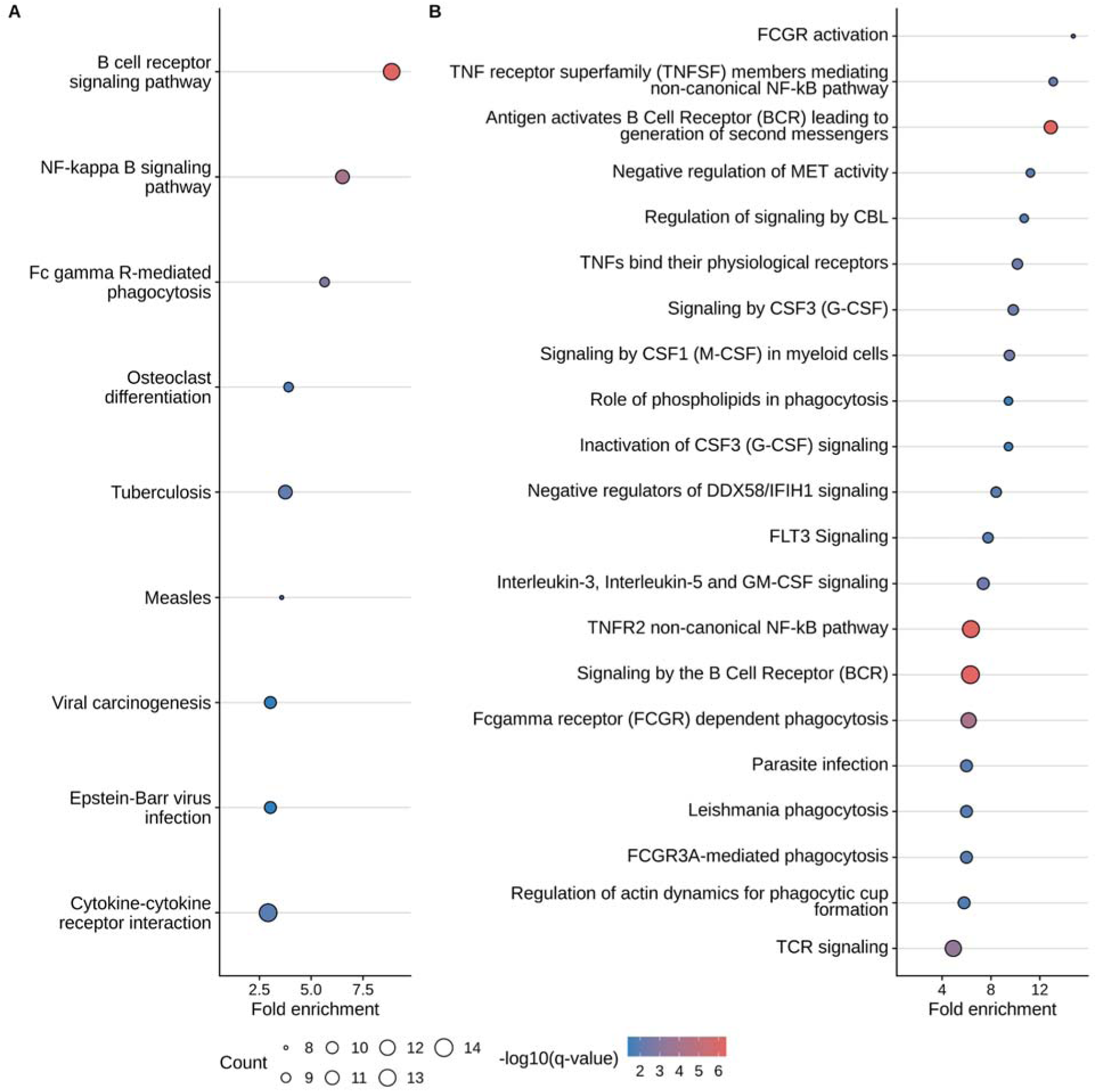
Significant (q-value < 0.05) pathways in (a) KEGG and (b) Reactome pathway analysis of the union of potential causal genes nominated for any isotype. The ‘fold enrichment’ is given by the ‘ratio of the frequency of input genes annotated in a term to the frequency of all genes annotated to that term’ (90).

In a few cases, we were able to relate the genes of isotype-specific associations to isotype-specific biology. We mapped the IgM-associated SNP rs614004 to *CMTM7*, which encodes a membrane protein important in the development of innate-like B1 cells which principally secrete constitutive, ‘natural’ IgM (53). We mapped associations for both IgA and IgG (i.e. class-switched isotypes), but not for IgM, to *TRAF3*, which encodes a regulator of class switch recombination (54). We linked the IgA-specific association on chromosome X to *C1GALT1C1*, essential for the O-glycosylation of IgA1 (the predominant IgA subclass in serum) and previously identified in a GWAS of galactose-deficient IgA1 (Gd-IgA1), an important IgA nephropathy endophenotype (55). *ST3GAL1*, another IgA-specific gene, encodes an enzyme also participating in O-glycosylation of IgA1. A recent GWAS of Gd-IgA1 also identified *DERL3*, a gene in which we report a missense variant associated with serum IgA and which participates in the processing of misfolded proteins (56). Despite studying a serum IgA phenotype, we also mapped an IgA-specific association to *LTBR*, essential for the development of Peyer’s patches, the site of secretory IgA production in the small intestine (57).

As antibodies are secreted by plasma cells and plasmablasts derived from B cells, we hypothesised that some signals may relate to B cell abundance. Whilst we could not find any sufficiently large GWAS of B cell count for use in colocalisation, total lymphocyte count has been studied in large cohorts, and we used one such GWAS as a proxy (27). We found a modest but significant genetic correlation between serum IgA and lymphocyte count (r_g_ = 0.09, p = 3 x 10^-4^). Whilst r_g_ estimates for IgM and IgG were not significant, we conducted colocalisation analyses for all three isotypes with lymphocyte counts as r_g_ had not proved a dispositive indicator of cross-isotype colocalisation.

We found 16 loci with moderate to strong evidence of a shared causal variant for serum antibody and lymphocyte count phenotypes (Supplementary Table 12): nine for IgA, six for IgM and one for IgG (minimum PPC = 0.69), including the above-mentioned signal near *GSDMB*, which colocalised with both IgA and IgM. However, the directions of effect were not consistent: higher immunoglobulin levels were associated with higher lymphocyte counts in eight cases and with lower lymphocyte counts in the other eight. This explains the lack of detectable genetic correlation, which is a genome-wide measure. Our interpretation of the ostensibly paradoxical relationship in the latter eight cases is hampered by the lack of B cell-specific data. Given lymphocyte count is the sum of B, T, and natural killer cell counts, an increased total lymphocyte count could arise from the production of more B cells, or fewer in the context of more T cells, or a change in T- or natural killer cell count absent any change in B-cell count.

All isotypes showed significant genetic correlation with two or more immune-mediated diseases (IMDs), amounting to nine significant estimates (Supplementary Table 13). These were positive with the exception of a negative genetic correlation between IgG and asthma (r_g_ = -0.1). We observed the strongest correlation between serum IgA and IgA nephropathy (r_g_ = 0.33), replicating published work (6). Inclusion of the *IGH*, *IGK*, and *IGL* loci made no nominally significant difference to antibody-immune disease genetic correlation estimates as assessed by z-test.

Several loci well-known for their association with IMDs colocalised with serum antibody signals, bolstering the evidence for the biological salience of the associations identified in our meta-analyses (Supplementary Figure 12). IgA associations signals at the *IL7R*, *INAVA*, *UBE2L3*, and *IL2RA* loci colocalised with three to five IMDs each (minimum PPC = 0.72) and, with one exception, SNP effects were positively correlated between the serum IgA and IMD phenotypes. We observed antibody-IMD signal colocalisation at the aforementioned *GSDMB* locus for eight isotype-IMD pairs (minimum PPC = 0.76), including all three studied isotypes. The IgM association signal centered upon the *PTPN22* missense variant rs2476601 colocalised strongly with those of rheumatoid arthritis, type 1 diabetes, Crohn’s disease, systemic lupus erythematosus, and hypothyroidism (minimum PPC = 1.0), IMDs whose association with this mutation is well-known (20,24,58). In general we observed that the direction of effect for serum antibody- and IMD-associated SNPs was neither consistently concordant nor discordant (Supplementary Figure 18). IgA nephropathy was exceptional in this regard: all four of its signals colocalising with serum IgA (minimum PPC = 0.86) showed positively correlated effects across the locus and at the lead SNP. Two of these signals mapped to the genes *TNFSF13* and *TNFRSF13B*, the APRIL-TACI receptor-ligand pair, and a third mapped to another TNF family member, *TNFSF8*; all three genes are implicated in B-cell biology.

68 of our associations were in or near known genes associated with inborn errors of immunity (IEIs); 32 of these associations were novel (Supplementary Table 14). With the exception of the more common antibody deficiencies, IEI phenotypes do not admit formal colocalisation analyses for want of summary statistics and evidence of common-variant architecture. For IgM, these associations included variants in *CARD11*, *LYN*, and *CIITA* as well as rs12554596, an intronic variant in and eQTL for *PAX5*, which encodes a transcription factor and ‘master regulator’ of B cell development (59). A single case of PAX5 deficiency is known, featuring B-cell lymphopaenia and hypogammaglobulinaemia (60). In two cases, variants associated with IgA and IgM appeared to represent distinct causal variants (on the basis of low LD and *coloc* favouring the distinct causal variant hypothesis) but both variants were linked to the same gene. IgM-associated rs290243 and IgA-associated rs4744011 are not in LD (r^2^=0.18), yet both belong to eQTL credible sets for *SYK* in NK cells and monocytes, respectively. *SYK* encodes a non-receptor tyrosine kinase which participates in BCR signalling (61) and in which gain-of-function mutations cause an autoinflammatory disorder (62). Similarly, rs6589225 and rs4938518 were associated with IgM and IgA respectively. Again, we observed these to be in low LD (r^2^=0.28) whilst belonging to eQTL credible sets for the same gene, *POU2AF1*. A loss-of-function mutation in *POU2AF1* has been observed to cause an autosomal recessive, CVID-like antibody deficiency, BOB1 deficiency, in one patient (63).

Other notable associations include IgA with rs1585213, an intronic variant in *NFKB1*, which encodes NF-κB, a critical immune regulator downstream of TLR and B-cell receptor (BCR) signalling and integral to development of the humoral response (64). Loss-of-function mutations in *NFKB1* have been identified as the most common cause of common variable immunodeficiency (65), an antibody deficiency characterised by IgG deficiency alongside IgA and/or IgM deficiency; IgA is absent in a majority of patients (66). Similarly, rs6890853 belongs to e-, s-, and pQTL credible sets for *IL7R*/*IL7R*, whose expression and signalling is important in the early stage of B-cell progenitor development. Loss-of-function mutations in *IL7R* cause the B cell-positive severe combined immunodeficiency IL7R deficiency (67).

Just as our pathway analyses linked association-related genes to B-cell biology, we could relate many individual novel antibody associations to the same. We found three novel IgA associations featuring missense variants in B cell-related genes. rs13388394 is located in *RASGRP3*, which encodes a guanine nucleotide exchange factor and RAS activator. Its murine orthologue *Rasgrp3* contributes to B-cell receptor-induced Ras activation and the deletion of *Rasgrp3* leads to deficient production of certain IgG subclasses (68). rs113495277 is a missense variant in *TRAF1*. TNF receptor associated factor 1 (TRAF1) forms multimeric complexes with TRAF2 which bind to the cytoplasmic domain of CD40 (the cell-surface receptor for the costimulatory ligand CD40L) upon ligation and activate the canonical NF-κB signalling pathway (69,70).

rs3177243 is a missense variant in *DERL3*, a gene recently associated with B cell activation and differentiation, and the unfolded protein response (71). Its association with the latter may explain its expression in plasma cells given their burden of secretory protein synthesis; rs3177243 is also associated with an IgG N-glycan phenotype (72) and non-albumin serum protein levels (73). We also identified a novel IgM-associated missense variant, rs11666267, in *NIBAN3*, which encodes B cell novel protein 1, a regulator of BCR signalling (74).

Other variants were linked to genes involved in B cell-related malignancies. This included two IgA-associated variants, rs55901664 and rs17505102, which were intronic to the genes *TBL1XR1* and *TP63*, respectively. Somatic mutations in *TBL1XR1* cause impaired plasma cell differentiation and a bias towards the production of immature memory B cells in aggressive B cell lymphomas (75), while *TP63* is known to participate in somatic gene fusion events with *TBL1XR1* in a small minority of diffuse large B-cell lymphomas (76).The IgG-associated variant rs184552867 is an intronic variant in *PRDM11* which has been identified as a tumour suppressor gene in B cells and which may be downregulated in diffuse large B-cell lymphoma (77).

## Discussion

The *IGH* and *IGK* loci encode immunoglobulin chains and are expected to be of central importance in a genetic analysis of serum antibody production. Indeed, germline variation within these loci has known effects upon the antibody repertoire and disease liability (78–81). However, the large and (within isotypes) discordant directions of effect we observed for serum IgG and serum IgA at the *IGH* locus suggest an artefactual epitope effect, particularly for the missense variant rs117775520.

As we cannot confidently report estimates obtained in these loci, we believe that the heritability estimates (and genetic correlation estimates derived from them) reported without the inclusion of the *IGH*, *IGK*, and *IGL* loci, and without the MHC, are more reliable. These estimates are also valuable in their own right as measures of the partial heritability of these traits attributable to genetic variation outside these important regions. We note that SumHer’s heritability estimation procedure depends upon a random effects model unsuited to isolated large effect sizes and the extreme LD seen in these regions. We saw an appreciable increase in estimated heritability with inclusion of the MHC, albeit calculated naively. Using long-read sequencing to improve resolution in the *IGH*, *IGK*, and *IGL* regions (82) together with more sophisticated analysis will be required for accurate determination of the heritability they explain in serum antibody levels. As we mapped strong signals to immunoglobulin heavy chain genes encoding distinct IgA and IgG subclasses (rs117775520 in particular is a missense variant in *IGHA1*), subclass-specific GWAS combined with long-read sequencing might provide the multimodal increase in resolution necessary to better understand these associations.

We detected positive phenotypic and genetic correlations between antibody isotypes but limited overlap in their individual association signals. This suggests that our cross-isotype genetic correlation estimates are driven by shared variant effects which do not achieve genome-wide significance despite our large sample sizes. Our list of associations thus remains an incomplete catalogue of the common-variant determinants of serum antibody production; we also report a SNP heritability estimate for serum IgA (0.18) which falls far short of the heritability estimate obtained from a past twin study (0.50) and the SNP heritability estimate (0.31) obtained in the same twin cohort (5). The effects we did detect reflect both known and novel biological mechanisms impacting immunoglobulin secretion, including B-cell progenitor development, antibody production and class switching, and the role of helper T cells in stimulating immunoglobulin production. Despite comparably-sized samples for IgG and IgM, we detected far fewer associations for the former and estimated its heritability to be around one third of the latter’s.

Outside of the *IGH*, *IGK*, and *IGL* loci, we do not believe technical effects can account for the lack of cross-isotype colocalisation. Each of our meta-analyses incorporated samples both from GWAS using immunoassays and those using aptamer-based assays. The composition with respect to assay type was similar for IgG and IgM: 79% and 84% came from aptamer-based studies for IgG and IgM, respectively, and 14% from immunoassay-based studies for both. The proportion of samples taken from immunoassay-based studies (35%) was higher for IgA, and that taken from aptamer-based studies (54%) lower, because of our inclusion of the immunoassay-based study by Liu et al. We believe it is unlikely that differences in sample composition between isotypes could introduce locus-specific biases in colocalisation outside of the immunoglobulin chain loci and in the face of the evidence of average genome-wide shared signal our genetic correlation estimates represent. More plausible is the possibility that the aggregation of IgG subclasses obscures subclass-specific colocalisations with IgM and IgA. Given recent evidence of IgG subclass-specific genetic architecture (80), a possible explanation for the smaller number of IgG associations we report is that the larger number of IgG subclasses (four compared with IgA’s two and IgM’s lack thereof) and our aggregation of them into one phenotype may have acted to dilute subclass-specific association signals. Future work could extend IgG and IgA subclass-specific GWAS to the same large scale as the meta-analyses we present here to clarify the subclasses driving the associations we report.

We found evidence of common-variant architecture shared between antibody phenotypes and immune-related traits. For the latter we observed multiple colocalisation signals indicative of shared causal variants in the absence of strong genetic correlation between lymphocyte count and the antibody phenotypes. Our expectation of straightforward concordance between antibody and lymphocyte count effect directions was refuted by the lack of a clear relationship. We observed the same inconsistency in effect direction across antibody signals colocalising with immune-mediated disease signals. These results suggest that immunoglobulin levels are not themselves on the causal path for any of these diseases, with the probable exception of IgA nephropathy (6). The shared associations instead reflect processes that lead to change in both immune-mediated disease risk and immunoglobulin levels. Our results also provide a cautionary tale for the use of genetic correlation more generally: weak or absent genetic correlation, as with lymphocyte counts, should not be taken as a dispositive indicator of genetic independence, and positive genetic correlation may be consistent with the presence of shared causal variants which nonetheless exert discordant effects on each phenotype.

We have presented an expanded view of the genetic architecture of serum antibody production. This view will be broadened still further by the adoption of modern high-throughput affinity-based proteomics techniques, such as the ‘SomaScan’ DNA aptamer-based method used to generate the Pietzner and Eldjarn datasets included in our meta-analyses (83). These methods allow many proteins to be assayed in parallel and should make the measurement of serum proteins in large, biobank-sized cohorts affordable (84). Investigation of the pattern of shared genetic effects on lymphocyte abundance and antibody production, and the seemingly paradoxical disagreement in the direction of variant effects upon them, was hindered by the lack of large, publicly available B and T cell-specific phenotype GWAS. Should such datasets become accessible, their use in colocalisation analysis should prove insightful as to the extent to which variants mediate their effect on serum antibody levels through proximal effects on lymphocyte abundance. The composite nature of the lymphocyte count phenotype in the available dataset prevented investigation of the causal effect of the abundance of B cells (or of relevant T helper cell subsets) on serum antibody production through Mendelian randomisation. B-cell lymphopaenia underlies hypo- and agammaglobulinaemia in a number of antibody deficiencies, such as Bruton’s agammaglobulinaemia and some CVID-like disorders such as TRNT1 deficiency (35). This suggests smaller genetic effects on B cell abundance mediated by common variants might also underlie variation in serum antibody production in the normophysiological setting.

We meta-analysed GWAS conducted almost exclusively in individuals of European ancestry, the sole exception being the trans-ancestry GWAS of serum IgA by Liu et al. We were thus not able to investigate the extent to which the genetic architecture of serum antibody production varies across ancestries or the replicability of our discoveries across populations. Whilst GWAS of antibody phenotypes in East Asian ancestries have been published (8,85), their summary statistics are not publicly available for meta-analysis, a common obstacle in GWAS meta-analysis (86).

As well as IMDs generally, our results are relevant to the study of the antibody deficiencies, particularly the two most common, selective IgA deficiency and common variable immunodeficiency (3). The genetic aetiology of these diseases remains obscure in most patients and there is evidence of a common-variant contribution to both (10,87,88). They are therefore good candidates for joint genetic analysis with the meta-analyses we publish here. Such an analysis could address the hypothesis of whether inheritance of a preponderance of common hypomorphic serum antibody-associated variants could account for clinical antibody deficiency in cases where a Mendelian pattern of disease or known monogenic causal variant are absent.

## Data and code availability

GWAS summary statistics have been deposited at the following Zenodo URL: https://doi.org/10.5281/zenodo.17010403.

We have made available the code used to produce the results presented here in the form of a *snakemake* (89) pipeline (doi: 10.5281/zenodo.18023612): https://github.com/twillis209/serum_ig_pipeline. The standalone *snakemake* pipeline used to process 1kGP data is also available: https://github.com/twillis209/1kGP_pipeline.

## Data Availability

All GWAS summary statistics produced in this work have been made publicly available through deposit at Zenodo (https://doi.org/10.5281/zenodo.17010403). Applications for access to individual-level genotype and phenotype data from the EPIC-Norfolk cohort can be made online (https://www.epic-norfolk.org.uk/for-researchers/data-sharing/data-requests/).

https://github.com/twillis209/serum_ig_pipeline

## Acknowledgements

We would like to acknowledge the participants of the Fenland Study (doi: 10.22025/2017.10.101.00001). We thank Christopher Benson, Debora Lucarelli, and Jing Hua Zhao for their contribution to the generation of data used in this paper. We wish to acknowledge all GWAS participants for their contribution to the data used herein. We also acknowledge the investigators who carried out these GWAS and made their summary statistics publicly available. We are grateful to all the participants of the EPIC-Norfolk study and to the many members of the study teams at the University of Cambridge who have enabled this research.

## Author contributions

Thomas W. Willis: Writing – review & editing, Writing – original draft, Visualization, Software, Methodology, Investigation, Formal analysis, Data curation, Conceptualization. Effrossyni Gkrania-Klotsas: Writing – review & editing, Methodology, Investigation, Funding acquisition, Data curation. Nicholas J. Wareham: Writing – review & editing, Funding acquisition, Data curation. Eoin F. McKinney: Writing – review & editing, Data curation. Paul A. Lyons: Writing – review & editing, Funding acquisition, Data curation. Kenneth G.C. Smith: Writing – review & editing, Funding acquisition. Chris Wallace: Writing – review & editing, Writing – original draft, Visualization, Supervision, Software, Methodology, Investigation, Funding acquisition, Formal analysis, Conceptualization.

## Bibliography

1. Hatagima A, Cabello PH, Krieger H. Causal analysis of the variability of IgA, IgG, and IgM immunoglobulin levels. Hum Biol. 1999;71(2):219–29.

2. Bruton OC. AGAMMAGLOBULINEMIA. Pediatrics. 1952 Jun;9(6):722–8.

3. Pan-Hammarström Q, Hammarström L. Antibody Deficiency Diseases. Eur J Immunol. 2008;38(2):327–33.

4. Yazdani R, Fekrvand S, Shahkarami S, Azizi G, Moazzami B, Abolhassani H, et al. The hyper IgM syndromes: Epidemiology, pathogenesis, clinical manifestations, diagnosis and management. Clin Immunol. 2019 Jan 1;198:19–30.

5. Viktorin A, Frankowiack M, Padyukov L, Chang Z, Melén E, Sääf A, et al. IgA measurements in over 12 000 Swedish twins reveal sex differential heritability and regulatory locus near CD30L. Hum Mol Genet. 2014 Aug 1;23(15):4177–84.

6. Liu L, Khan A, Sanchez-Rodriguez E, Zanoni F, Li Y, Steers N, et al. Genetic Regulation of Serum IgA Levels and Susceptibility to Common Immune, Infectious, Kidney, and Cardio-Metabolic Traits. Nat Commun. 2022 Nov;13(1):6859.

7. Bian S, Guo X, Yang X, Wei Y, Yang Z, Cheng S, et al. Genetic determinants of IgG antibody response to COVID-19 vaccination. Am J Hum Genet. 2024 Jan 4;111(1):181–99.

8. Yang M, Wu Y, Lu Y, Liu C, Sun J, Liao M, et al. Genome-wide scan identifies variant in TNFSF13 associated with serum IgM in a healthy Chinese male population. PLoS One. 2012 Oct 31;7(10):e47990.

9. Kim JH, Cheong HS, Park JS, Jang AS, Uh ST, Kim YH, et al. A genome-wide association study of total serum and mite-specific IgEs in asthma patients. PLoS One. 2013 Aug 13;8(8):e71958.

10. Willis TW, Gkrania-Klotsas E, Wareham NJ, McKinney EF, Lyons PA, Smith KGC, et al. Leveraging pleiotropy identifies common-variant associations with selective IgA deficiency. Clin Immunol. 2024 Nov 1;268(110356):110356.

11. Dennis JK, Sealock JM, Straub P, Lee YH, Hucks D, Actkins K’era, et al. Clinical Laboratory Test-Wide Association Scan of Polygenic Scores Identifies Biomarkers of Complex Disease. Genome Med. 2021 Jan;13(1):6.

12. Scepanovic P, Alanio C, Hammer C, Hodel F, Bergstedt J, Patin E, et al. Human genetic variants and age are the strongest predictors of humoral immune responses to common pathogens and vaccines. Genome Med. 2018 Jul 27;10(1):59.

13. Eldjarn GH, Ferkingstad E, Lund SH, Helgason H, Magnusson OT, Gunnarsdottir K, et al. Large-scale plasma proteomics comparisons through genetics and disease associations. Nature. 2023 Oct 4;622(7982):348–58.

14. Pietzner M, Wheeler E, Carrasco-Zanini J, Cortes A, Koprulu M, Wörheide MA, et al. Mapping the proteo-genomic convergence of human diseases. Science. 2021 Nov 12;374(6569):eabj1541.

15. Jonsson S, Sveinbjornsson G, de Lapuente Portilla AL, Swaminathan B, Plomp R, Dekkers G, et al. Identification of Sequence Variants Influencing Immunoglobulin Levels. Nat Genet. 2017 Aug;49(8):1182–91.

16. Cordell HJ, Fryett JJ, Ueno K, Darlay R, Aiba Y, Hitomi Y, et al. An International Genome-Wide Meta-Analysis of Primary Biliary Cholangitis: Novel Risk Loci and Candidate Drugs. J Hepatol. 2021 Sep;75(3):572–81.

17. Ji SG, Juran BD, Mucha S, Folseraas T, Jostins L, Melum E, et al. Genome-Wide Association Study of Primary Sclerosing Cholangitis Identifies New Risk Loci and Quantifies the Genetic Relationship with Inflammatory Bowel Disease. Nat Genet. 2017 Feb;49(2):269–73.

18. Ishigaki K, Sakaue S, Terao C, Luo Y, Sonehara K, Yamaguchi K, et al. Multi-Ancestry Genome-Wide Association Analyses Identify Novel Genetic Mechanisms in Rheumatoid Arthritis. Nat Genet. 2022 Nov;54(11):1640–51.

19. Bentham J, Morris DL, Cunninghame Graham DS, Pinder CL, Tombleson P, Behrens TW, et al. Genetic Association Analyses Implicate Aberrant Regulation of Innate and Adaptive Immunity Genes in the Pathogenesis of Systemic Lupus Erythematosus. Nat Genet. 2015 Dec;47(12):1457–64.

20. de Lange KM, Moutsianas L, Lee JC, Lamb CA, Luo Y, Kennedy NA, et al. Genome-Wide Association Study Implicates Immune Activation of Multiple Integrin Genes in Inflammatory Bowel Disease. Nat Genet. 2017 Feb;49(2):256–61.

21. Chiou J, Geusz RJ, Okino ML, Han JY, Miller M, Melton R, et al. Interpreting Type 1 Diabetes Risk with Genetics and Single-Cell Epigenomics. Nature. 2021 Jun;594(7863):398–402.

22. INTERNATIONAL MULTIPLE SCLEROSIS GENETICS CONSORTIUM. Multiple Sclerosis Genomic Map Implicates Peripheral Immune Cells and Microglia in Susceptibility. Science. 2019 Sep;365(6460):eaav7188.

23. Pasanen A, Sliz E, Huilaja L, FinnGen, Estonian Biobank Research Team, Reimann E, et al. Identifying atopic dermatitis risk loci in 1,094,060 individuals with subanalysis of disease severity and onset. J Invest Dermatol. 2024 Nov;144(11):2417–25.

24. Figuerêdo J, Krebs K, Pujol-Gualdo N, Haller T, Võsa U, Volke V, et al. Uncovering the shared genetic components of thyroid disorders and reproductive health. Eur J Endocrinol. 2024 Aug 5;191(2):211–22.

25. Trynka G, Hunt KA, Bockett NA, Romanos J, Mistry V, Szperl A, et al. Dense Genotyping Identifies and Localizes Multiple Common and Rare Variant Association Signals in Celiac Disease. Nat Genet. 2011 Dec;43(12):1193–201.

26. Kiryluk K, Sanchez-Rodriguez E, Zhou XJ, Zanoni F, Liu L, Mladkova N, et al. Genome-wide association analyses define pathogenic signaling pathways and prioritize drug targets for IgA nephropathy. Nat Genet. 2023 Jul;55(7):1091–105.

27. Chen MH, Raffield LM, Mousas A, Sakaue S, Huffman JE, Moscati A, et al. Trans-Ethnic and Ancestry-Specific Blood-Cell Genetics in 746,667 Individuals from 5 Global Populations. Cell. 2020 Sep;182(5):1198–213.e14.

28. Cerezo M, Sollis E, Ji Y, Lewis E, Abid A, Bircan KO, et al. The NHGRI-EBI GWAS Catalog: standards for reusability, sustainability and diversity. Nucleic Acids Res. 2025 Jan 6;53(D1):D998–1005.

29. Chang CC, Chow CC, Tellier LC, Vattikuti S, Purcell SM, Lee JJ. Second-Generation PLINK: Rising to the Challenge of Larger and Richer Datasets. Gigascience. 2015 Dec;4(1):s13742–015 – 0047–8.

30. Purcell S, Chang C. PLINK 2.0.

31. Singh A, Southam L, Hatzikotoulas K, Rayner NW, Suzuki K, Taylor HJ, et al. Correcting for genomic inflation leads to loss of power in large-scale genome-wide association study meta-analysis. Genet Epidemiol. 2025 Sep 6;49(6):e70016.

32. Lewis MJ, Wang S. locuszoomr: an R package for visualizing publication-ready regional gene locus plots. Bioinform Adv. 2024 Dec 26;5(1):vbaf006.

33. Machiela MJ, Chanock SJ. LDlink: a web-based application for exploring population-specific haplotype structure and linking correlated alleles of possible functional variants. Bioinformatics. 2015 Nov 1;31(21):3555–7.

34. Gudjonsson A, Gudmundsdottir V, Axelsson GT, Gudmundsson EF, Jonsson BG, Launer LJ, et al. A genome-wide association study of serum proteins reveals shared loci with common diseases. Nat Commun. 2022 Jan 25;13(1):480.

35. Poli MC, Aksentijevich I, Bousfiha AA, Cunningham-Rundles C, Hambleton S, Klein C, et al. Human inborn errors of immunity: 2024 update on the classification from the International Union of Immunological Societies Expert Committee. Journal of Human Immunity [Internet]. 2025 May 5 [cited 2025 Aug 10];1(1). Available from: 10.70962/jhi.20250003

36. Durinck S, Spellman PT, Birney E, Huber W. Mapping identifiers for the integration of genomic datasets with the R/Bioconductor package biomaRt. Nat Protoc. 2009 Jul 23;4(8):1184–91.

37. Durinck S, Moreau Y, Kasprzyk A, Davis S, De Moor B, Brazma A, et al. BioMart and Bioconductor: a powerful link between biological databases and microarray data analysis. Bioinformatics. 2005 Aug 15;21(16):3439–40.

38. Speed D, Balding DJ. SumHer Better Estimates the SNP Heritability of Complex Traits from Summary Statistics. Nat Genet. 2019 Feb;51(2):277–84.

39. A Global Reference for Human Genetic Variation. Nature. 2015 Oct;526(7571):68–74.

40. Byrska-Bishop M, Evani US, Zhao X, Basile AO, Abel HJ, Regier AA, et al. High Coverage Whole Genome Sequencing of the Expanded 1000 Genomes Project Cohort Including 602 Trios. bioRxiv. 2021 Feb;2021.02.06.430068.

41. Buniello A, Suveges D, Cruz-Castillo C, Llinares MB, Cornu H, Lopez I, et al. Open Targets Platform: facilitating therapeutic hypotheses building in drug discovery. Nucleic Acids Res. 2025 Jan 6;53(D1):D1467–75.

42. Dyer SC, Austine-Orimoloye O, Azov AG, Barba M, Barnes I, Barrera-Enriquez VP, et al. Ensembl 2025. Nucleic Acids Res. 2025 Jan 6;53(D1):D948–57.

43. Yu G. Thirteen years of clusterProfiler. Innovation (Camb). 2024 Nov 4;5(6):100722.

44. Yu G, He QY. ReactomePA: an R/Bioconductor package for reactome pathway analysis and visualization. Mol Biosyst. 2016 Feb 26;12(2):477–9.

45. Giambartolomei C, Vukcevic D, Schadt EE, Franke L, Hingorani AD, Wallace C, et al. Bayesian Test for Colocalisation between Pairs of Genetic Association Studies Using Summary Statistics. PLoS Genet. 2014 May;10(5):e1004383.

46. Wakefield J. Bayes factors for genome-wide association studies: comparison with P-values. Genet Epidemiol. 2009 Jan 1;33(1):79–86.

47. Kanai M, Elzur R, Zhou W, Global Biobank Meta-analysis Initiative, Daly MJ, Finucane HK. Meta-analysis fine-mapping is often miscalibrated at single-variant resolution. Cell Genom. 2022 Dec 14;2(12):100210.

48. Jefferis R, Lefranc MP. Human immunoglobulin allotypes: possible implications for immunogenicity. MAbs. 2009 Jul;1(4):332–8.

49. Purcell RA, Aurelia LC, Esterbauer R, Allen LF, Bond KA, Williamson DA, et al. Immunoglobulin G genetic variation can confound assessment of antibody levels via altered binding to detection reagents. Clin Transl Immunology. 2024 Feb 29;13(3):e1494.

50. Li X, Bu X, Lu B, Avraham H, Flavell RA, Lim B. The hematopoiesis-specific GTP-binding protein RhoH is GTPase deficient and modulates activities of other Rho GTPases by an inhibitory function. Mol Cell Biol. 2002 Feb;22(4):1158–71.

51. Crequer A, Troeger A, Patin E, Ma CS, Picard C, Pedergnana V, et al. Human RHOH deficiency causes T cell defects and susceptibility to EV-HPV infections. J Clin Invest. 2012 Sep 4;122(9):3239–47.

52. Dorn T. Analysis of RhoH function in vivo [Internet] [Doctoral dissertation]. [Ludwig-Maximiliansuniversitaet Muenchen]: Ludwig-Maximiliansuniversitaet Muenchen; 2007. Available from: https://edoc.ub.uni-muenchen.de/7457/1/Dorn_Tatjana.pdf

53. Liu Z, Liu Y, Li T, Wang P, Mo X, Lv P, et al. CMTM7 plays key roles in TLR-induced plasma cell differentiation and p38 activation in murine B-1 B cells. Eur J Immunol. 2020 Jun 1;50(6):809–21.

54. Chen Z, Krinsky A, Woolaver RA, Wang X, Chen SMY, Popolizio V, et al. TRAF3 acts as a checkpoint of B cell receptor signaling to control antibody class switch recombination and anergy. J Immunol. 2020 Aug 1;205(3):830–41.

55. Kiryluk K, Li Y, Moldoveanu Z, Suzuki H, Reily C, Hou P, et al. GWAS for serum galactose-deficient IgA1 implicates critical genes of the O-glycosylation pathway. PLoS Genet. 2017 Feb 10;13(2):e1006609.

56. Liu L, Shen A, Taylor AD, Atlas Khan, Reily C, Hall SD, et al. Genome-wide association study (GWAS) of serum galactose-deficient IgA1 uncovers shared genetic determinants with IgAN: Th-po0633. J Am Soc Nephrol. 2025 Oct;36(10S):10.1681/ASN.2025pyp4sfbe.

57. Fütterer A, Mink K, Luz A, Kosco-Vilbois MH, Pfeffer K. The lymphotoxin β receptor controls organogenesis and affinity maturation in peripheral lymphoid tissues. Immunity. 1998 Jul;9(1):59–70.

58. Eyre S, Hinks A, Bowes J, Flynn E, Martin P, Wilson AG, et al. Overlapping Genetic Susceptibility Variants between Three Autoimmune Disorders: Rheumatoid Arthritis, Type 1 Diabetes and Coeliac Disease. Arthritis Res Ther. 2010 Sep;12(5):R175.

59. Medvedovic J, Ebert A, Tagoh H, Busslinger M. Pax5: a master regulator of B cell development and leukemogenesis. Adv Immunol. 2011;111:179–206.

60. Kaiser FMP, Gruenbacher S, Oyaga MR, Nio E, Jaritz M, Sun Q, et al. Biallelic PAX5 mutations cause hypogammaglobulinemia, sensorimotor deficits, and autism spectrum disorder. J Exp Med [Internet]. 2022 Sep 5 [cited 2025 Feb 21];219(9). Available from: https://rupress.org/jem/article-pdf/219/9/e20220498/1916562/jem_20220498.pdf

61. Ackermann JA, Nys J, Schweighoffer E, McCleary S, Smithers N, Tybulewicz VLJ. Syk tyrosine kinase is critical for B cell antibody responses and memory B cell survival. J Immunol. 2015 May 15;194(10):4650–6.

62. Wang L, Aschenbrenner D, Zeng Z, Cao X, Mayr D, Mehta M, et al. Gain-of-function variants in SYK cause immune dysregulation and systemic inflammation in humans and mice. Nat Genet. 2021 Apr 29;53(4):500–10.

63. Kury P, Staniek J, Wegehaupt O, Janowska I, Eckenweiler M, Korinthenberg R, et al. Agammaglobulinemia with Normal B-cell Numbers in a Patient Lacking Bob1. J Allergy Clin Immunol. 2021 May;147(5):1977–80.

64. Guldenpfennig C, Teixeiro E, Daniels M. NF-kB’s contribution to B cell fate decisions. Front Immunol. 2023 Jul 18;14:1214095.

65. Tuijnenburg P, Lango Allen H, Burns SO, Greene D, Jansen MH, Staples E, et al. Loss-of-Function Nuclear Factor κB Subunit 1 (NFKB1) Variants Are the Most Common Monogenic Cause of Common Variable Immunodeficiency in Europeans. J Allergy Clin Immunol. 2018 Oct;142(4):1285–96.

66. Bonilla FA, Barlan I, Chapel H, Costa-Carvalho BT, Cunningham-Rundles C, de la Morena MT, et al. International Consensus Document (ICON): Common Variable Immunodeficiency Disorders. J Allergy Clin Immunol Pract. 2016 Jan;4(1):38–59.

67. Kaiser FMP, Janowska I, Menafra R, de Gier M, Korzhenevich J, Pico-Knijnenburg I, et al. IL-7 receptor signaling drives human B-cell progenitor differentiation and expansion. Blood. 2023 Sep 28;142(13):1113–30.

68. Coughlin JJ, Stang SL, Dower NA, Stone JC. RasGRP1 and RasGRP3 regulate B cell proliferation by facilitating B cell receptor-Ras signaling. J Immunol. 2005 Dec 1;175(11):7179–84.

69. Elgueta R, Benson MJ, de Vries VC, Wasiuk A, Guo Y, Noelle RJ. Molecular mechanism and function of CD40/CD40L engagement in the immune system. Immunol Rev. 2009 May;229(1):152–72.

70. Fotin-Mleczek M, Henkler F, Hausser A, Glauner H, Samel D, Graness A, et al. Tumor necrosis factor receptor-associated factor (TRAF) 1 regulates CD40-induced TRAF2-mediated NF-kappaB activation. J Biol Chem. 2004 Jan 2;279(1):677–85.

71. Lin L, Lin G, Lin H, Chen L, Chen X, Lin Q, et al. Integrated profiling of endoplasmic reticulum stress-related DERL3 in the prognostic and immune features of lung adenocarcinoma. Front Immunol. 2022 Oct 7;13:906420.

72. Landini A, Trbojević-Akmačić I, Navarro P, Tsepilov YA, Sharapov SZ, Vučković F, et al. Genetic regulation of post-translational modification of two distinct proteins. Nat Commun. 2022 Mar 24;13(1):1586.

73. Sinnott-Armstrong N, Tanigawa Y, Amar D, Mars N, Benner C, Aguirre M, et al. Genetics of 35 blood and urine biomarkers in the UK Biobank. Nat Genet. 2021 Feb 18;53(2):185–94.

74. Hong R, Lai N, Ouchida R, Xiong E, Zhou Y, Min Q, et al. The B cell novel protein 1 (BCNP1) regulates BCR signaling and B cell apoptosis. Eur J Immunol. 2019 Jun;49(6):911–7.

75. Venturutti L, Teater M, Zhai A, Chadburn A, Babiker L, Kim D, et al. TBL1XR1 mutations drive extranodal lymphoma by inducing a pro-tumorigenic memory fate. Cell. 2020 Jul 23;182(2):297–316.e27.

76. Scott DW, Mungall KL, Ben-Neriah S, Rogic S, Morin RD, Slack GW, et al. TBL1XR1/TP63: a novel recurrent gene fusion in B-cell non-Hodgkin lymphoma. Blood. 2012 May 24;119(21):4949–52.

77. Asmar F, Fog CK, Jensen K, Jacobsen L, Ralfkiær E, Geisler CH, et al. PRDM11 is a novel putative tumor suppressor in aggressive B Cell Lymphoma. Blood. 2010 Nov 19;116(21):1002–1002.

78. Rodriguez OL, Safonova Y, Silver CA, Shields K, Gibson WS, Kos JT, et al. Genetic variation in the immunoglobulin heavy chain locus shapes the human antibody repertoire. Nat Commun. 2023 Jul 21;14(1):4419.

79. Engelbrecht E, Rodriguez OL, Lees W, Vanwinkle Z, Shields K, Schultze S, et al. Germline polymorphisms in the immunoglobulin kappa and lambda loci underpinning antibody light chain repertoire variability. Nat Commun. 2025 Nov 28;16(1):11707.

80. Olafsdottir TA, Thorleifsson G, Lopez de Lapuente Portilla A, Jonsson S, Stefansdottir L, Niroula A, et al. Sequence variants influencing the regulation of serum IgG subclass levels. Nat Commun. 2024 Sep 14;15(1):8054.

81. Corcoran M, Narang S, Kaduk M, Chernyshev M, Färnert A, Sundling C, et al. Human IGH germline gene diversity and allele frequencies in 2486 individuals from 25 global populations delineated by ultra-high throughput genotyping [Internet]. bioRxiv. bioRxiv; 2025 [cited 2026 Jan 8]. p. 2025.08.06.668935. Available from: https://www.biorxiv.org/content/10.1101/2025.08.06.668935v1.abstract

82. Watson CT, Collins AM, Ohlin M, Heather JM, Peres A, Lees WD, et al. Building immunoglobulin and T cell receptor gene databases for the future. Immunoinformatics (Amst). 2025 Sep 1;19(100059):100059.

83. Gold L, Walker JJ, Wilcox SK, Williams S. Advances in human proteomics at high scale with the SOMAscan proteomics platform. N Biotechnol. 2012 Jun 15;29(5):543–9.

84. Largest proteome study enlists 14 biopharmas. Nat Biotechnol. 2025 Feb 14;43(2):153.

85. Kanai M, Akiyama M, Takahashi A, Matoba N, Momozawa Y, Ikeda M, et al. Genetic analysis of quantitative traits in the Japanese population links cell types to complex human diseases. Nat Genet. 2018 Mar 5;50(3):390–400.

86. Reales G, Wallace C. Sharing GWAS Summary Statistics Results in More Citations. Communications Biology. 2023 Jan;6(1):1–6.

87. Li J, Jørgensen SF, Maggadottir SM, Bakay M, Warnatz K, Glessner J, et al. Association of CLEC16A with Human Common Variable Immunodeficiency Disorder and Role in Murine B Cells. Nat Commun. 2015 Apr;6:6804.

88. Bronson PG, Chang D, Bhangale T, Seldin MF, Ferreira RC, Urcelay E, et al. Common Variants at PVT1, ATG13-AMBRA1, AHI1 and CLEC16A Are Associated with Selective IgA Deficiency. Nat Genet. 2016 Nov;48(11):1425–9.

89. Mölder F, Jablonski KP, Letcher B, Hall MB, Tomkins-Tinch CH, Sochat V, et al. Sustainable Data Analysis with Snakemake. F1000Res [Internet]. 2021 Apr [cited 2022 Sep 27];10(33). Available from: 10.12688/f1000research.29032.2

90. Wu T, Hu E, Xu S, Chen M, Guo P, Dai Z, et al. clusterProfiler 4.0: A universal enrichment tool for interpreting omics data. Innovation (Camb). 2021 Aug 28;2(3):100141.

