## Supplementary Figures and Table Captions for "Large-scale GWAS meta-analysis of serum antibody levels in healthy individuals reveals distinct genetic architectures"

### Supplementary Table Legends

**Supplementary Table 1**. The serum antibody GWAS subject to meta-analysis in this work.

**Supplementary Table 2**. The non-antibody GWAS used in this work. Where studies are not available on the EBI GWAS Catalog, we give publicly accessible URLs where available.

**Supplementary Table 3**. The transformation applied to each dataset included in our meta-analysis in its original publication.

**Supplementary Table 4.** Lead SNPs from genome-wide significant associations in the *IGH*, *IGK*, and *IGL* loci in the serum antibody meta-analyses and GWAS included in the meta-analyses. 'Gene(s)' gives the gene(s) with the most evidence linking it/them to the association signal; the procedure by which we assigned genes to variants, and identified quantitative trait loci (QTL) and missense variants is described in Methods. ‘Missense gene’ reports the gene (if any) in which the lead SNP was identified to be a missense variant. ‘Q’ is Cochran’s Q test statistic for which the degrees of freedom and p-value are also given. ‘I2’ is the I^2^ statistic.

**Supplementary Table 5.** Lead SNPs from genome-wide significant associations in the serum IgG meta-analysis. ‘MAF’ is the minor allele frequency computed from data in the component GWAS where this information was provided in the original studies. 'Gene(s)' gives the gene(s) with the most evidence linking it/them to the association signal; the procedure by which we assigned genes to variants, and identified quantitative trait loci (QTL) and missense variants is described in Methods. ‘Missense gene’ reports the gene (if any) in which the lead SNP was identified to be a missense variant. ‘Q’ is Cochran’s Q test statistic for which the degrees of freedom and p-value are also given. ‘I2’ is the I^2^ statistic.

**Supplementary Table 6.** Lead SNPs from genome-wide significant associations in the serum IgM meta-analysis. ‘MAF’ is the minor allele frequency computed from data in the component GWAS where this information was provided in the original studies. 'Gene(s)' gives the gene(s) with the most evidence linking it/them to the association signal; the procedure by which we assigned genes to variants, and identified quantitative trait loci (QTL) and missense variants is described in Methods. ‘Missense gene’ reports the gene (if any) in which the lead SNP was identified to be a missense variant. ‘Q’ is Cochran’s Q test statistic for which the degrees of freedom and p-value are also given. ‘I2’ is the I^2^ statistic.

**Supplementary Table 7.** Lead SNPs from genome-wide significant associations in the serum IgA meta-analysis. ‘MAF’ is the minor allele frequency computed from data in the component GWAS where this information was provided in the original studies. 'Gene(s)' gives the gene(s) with the most evidence linking it/them to the association signal; the procedure by which we assigned genes to variants, and identified quantitative trait loci (QTL) and missense variants is described in Methods. ‘Missense gene’ reports the gene (if any) in which the lead SNP was identified to be a missense variant. ‘Q’ is Cochran’s Q test statistic for which the degrees of freedom and p-value are also given. ‘I2’ is the I^2^ statistic.

**Supplementary Table 8.** Heritability estimates for serum antibody phenotypes. The ‘MHC’ column indicates whether variants lying within the major histocompatibility complex were included in the dataset for heritability estimation. The ‘IGHKL’ indicates inclusion or exclusion of variants lying in the *IGH*, *IGK*, and *IGL* loci.

**Supplementary Table 9.** Cross-isotype phenotypic correlations in the EPIC cohort. ‘Scale’ indicates whether untransformed (‘raw’) or log-transformed antibody concentrations were used to compute correlations. ‘95% CI’ is 95% confidence interval.

**Supplementary Table 10**. Estimates of the genetic correlation between serum antibody isotype levels. p-values were calculated from a chi-squared test of non-zero genetic correlation. The ‘MHC’ and ‘IGHKL’ columns indicate inclusion of the MHC and *IGH*, *IGK*, and *IGL* loci in the datasets used to produce each estimate.

**Supplementary Table 11.** The results of the antibody-antibody colocalisation analyses. ‘No. of SNPs’ is the number of SNPs included in the analysis. Columns ‘PP.H0.abf’ to ‘PP.H4.abf’ give the posterior probability of the *coloc* hypotheses H_0_ to H_4_. ‘Filtered’ indicates whether the analysis was performed on data subject to the filtering process described in the colocalisation section of Methods. ‘Min. locus p-value’ is the minimum p-value across all SNPs considered in the colocalisation analysis. The ‘Pearson correlation’ column gives the Pearson correlation of Z-scores between the antibody phenotypes. The ‘Effect ratio’ columns give the ratio of the first and second isotype’s effect estimates at the lead SNP of the former and latter, respectively; where an effect ratio is absent, this is due to the absence of one phenotype’s lead SNP in the other phenotype’s dataset. ‘r2’ gives the squared correlation coefficient of genotypes between the lead SNPs of the two isotypes.

**Supplementary Table 12.** The results of the antibody-immune trait (‘non-Ig trait’) colocalisation analyses. ‘No. of SNPs’ is the number of SNPs included in the analysis. Columns ‘PP.H0.abf’ to ‘PP.H4.abf’ give the posterior probability of the *coloc* hypotheses H_0_ to H_4_. ‘Min. locus p-value’ is the minimum p-value across all SNPs considered in the colocalisation analysis. The ‘Pearson correlation’ column gives the Pearson correlation of Z-scores between the antibody and non-antibody phenotypes. The ‘Effect ratio’ columns give the ratio of the antibody and immune trait effect estimates at the lead SNP of the former and latter, respectively; where an effect ratio is absent, this is due to the absence of one phenotype’s lead SNP in the other phenotype’s dataset. ‘r2’ gives the squared correlation coefficient of genotypes between the lead SNPs of the antibody and immune phenotypes.

**Supplementary Table 13**. Estimates of the genetic correlation between antibody and immune phenotypes. We give estimates, their standard errors, and their p-values for datasets with and without inclusion of the *IGH*, *IGK*, and *IGL* loci. p-values were calculated from a chi-squared test of non-zero genetic correlation. The ‘FDR’ column gives the minimum false discovery rate at which the null of no genetic correlation would be rejected by the Benjamini-Hochberg procedure applied to the p-values obtained without inclusion of the *IGH*, *IGK*, and *IGL* loci.

**Supplementary Table 14**. Inborn errors of immunity (IEI) with causal variants occurring in genes within 200kb of the lead SNPs for antibody associations identified in this work. 'Novel' indicates whether an association with serum antibody has previously been reported for a variant. ‘Distance to gene’ gives the distance in base pairs from the antibody lead SNP to the IEI gene.

### Supplementary Figures

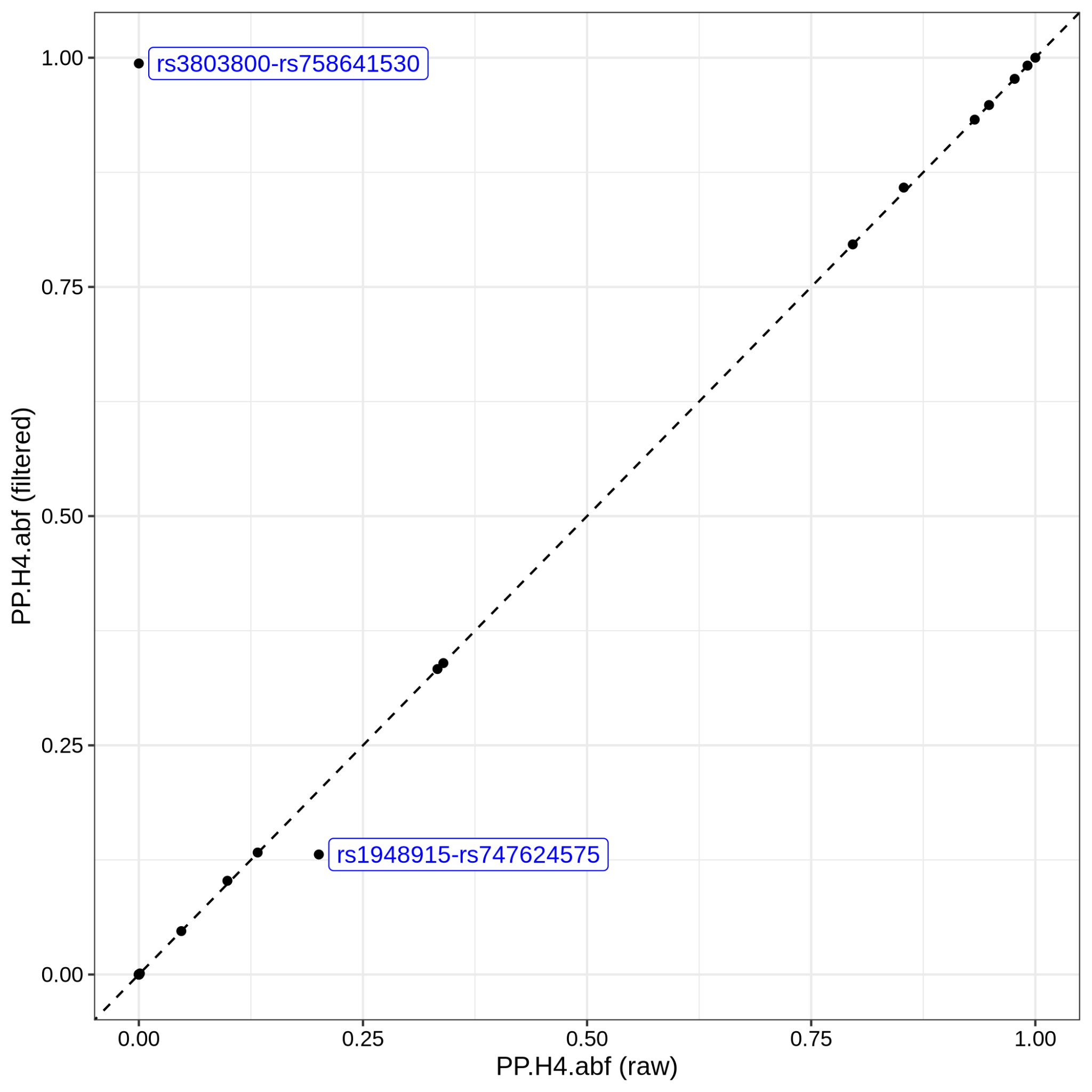

**Supplementary Figure 1.** The posterior probability of the ‘H4’ hypothesis of a shared causal variant (‘PP.H4.abf’) in *coloc* in antibody-antibody colocalisation analyses with (‘filtered’) and without (‘raw’) filtering of SNPs by sample fraction. The dashed line indicates y = x. The two labelled points are those lying an appreciable distance from the line y = x.

**Supplementary Figure 2**. Colocalisation analysis of the IgA and IgG association signals on chromosome 17 in the interval 6.9Mb to 7.8Mb without filtering. The left half of the Figure depicts meta-analytic p-values for the variants in the interval for each phenotype; lead SNPs for each phenotype’s association are coloured red and labelled. SNPs with p-values below 5x10^-8^ are coloured blue. The panel below the Manhattan plots depicts the genes located within the same genomic interval. The top panel on the right half of the Figure depicts SNP Z-scores for each phenotype. The dashed blue lines give the threshold for genome-wide significance on the Z-score scale. The table in the lower half gives statistics relating to the colocalisation analysis. ‘min(p)’ is the smallest p-value in the genomic interval subject to colocalisation analysis. The ‘PP.Hx.abf’ statistics give the posterior probability of the colocalisation hypotheses H_0_ to H_4_.
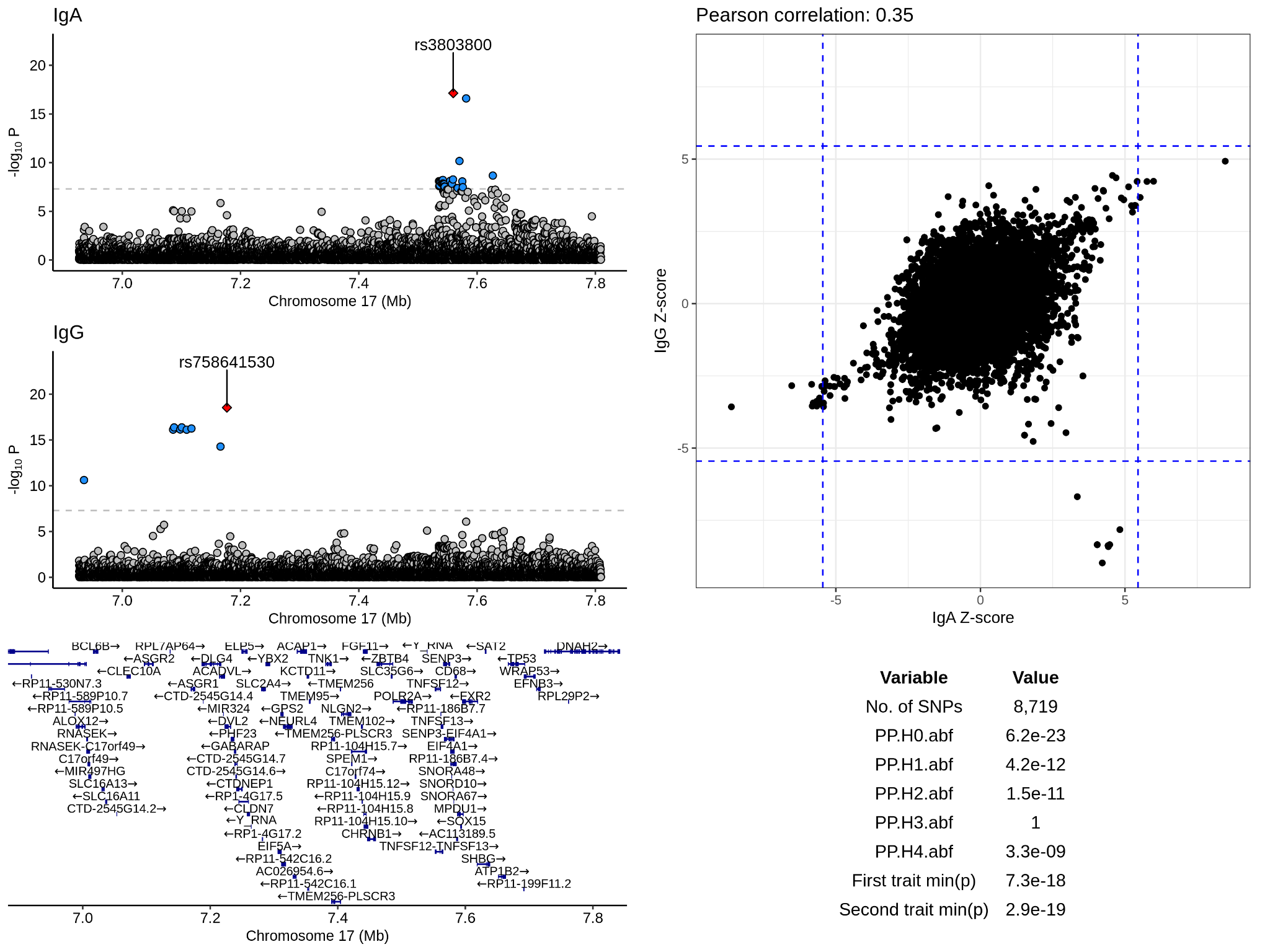

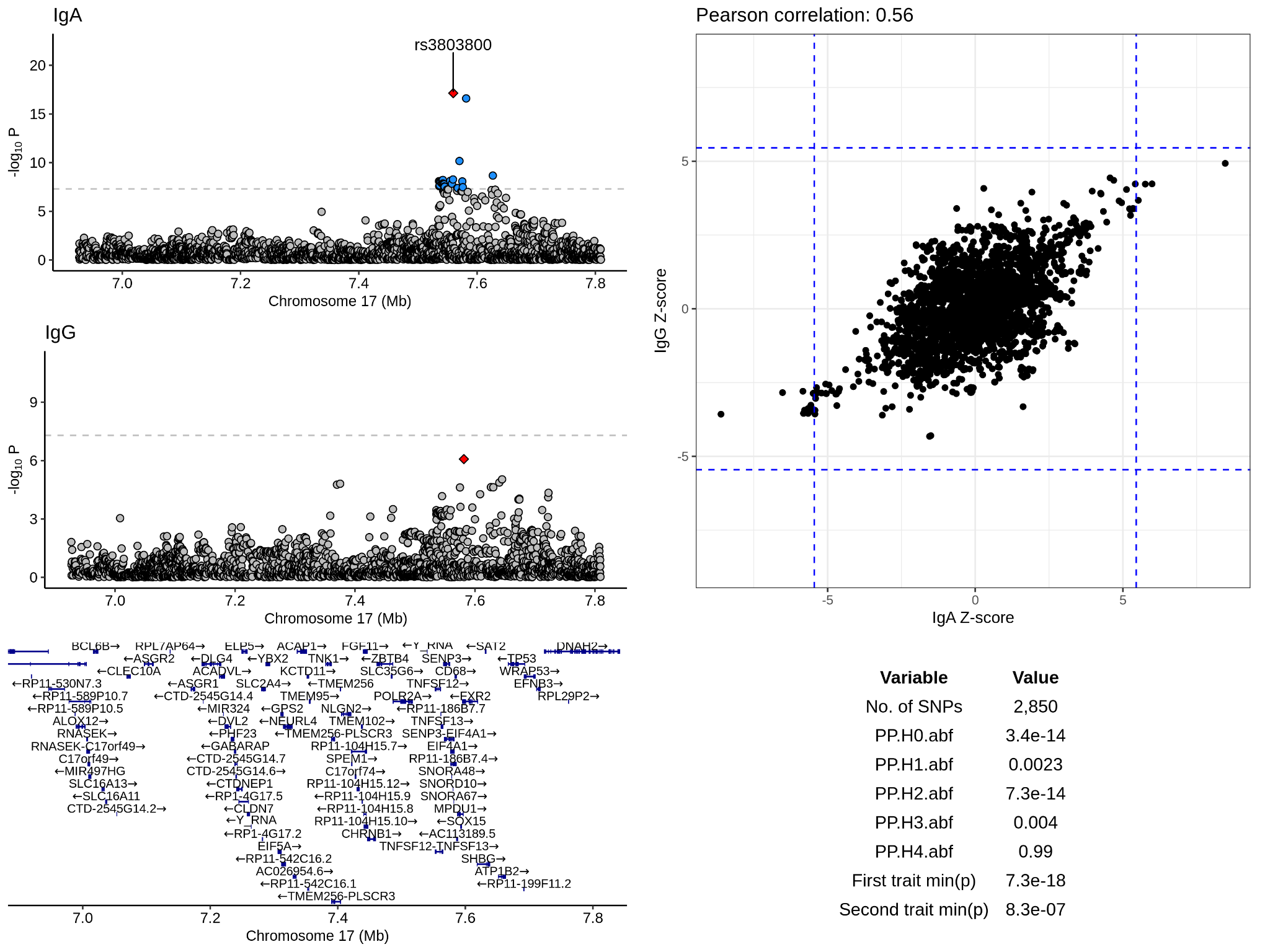

**Supplementary Figure 3**. Colocalisation analysis of the IgA and IgG association signals on chromosome 17 in the interval 6.9Mb to 7.8Mb with filtering. The left half of the Figure depicts meta-analytic p-values for the variants in the interval for each phenotype; lead SNPs for each phenotype’s association are coloured red and labelled. SNPs with p-values below 5x10^-8^ are coloured blue. The panel below the Manhattan plots depicts the genes located within the same genomic interval. The top panel on the right half of the Figure depicts SNP Z-scores for each phenotype. The dashed blue lines give the threshold for genome-wide significance on the Z-score scale. The table in the lower half gives statistics relating to the colocalisation analysis. ‘min(p)’ is the smallest p-value in the genomic interval subject to colocalisation analysis. The ‘PP.Hx.abf’ statistics give the posterior probability of the colocalisation hypotheses H_0_ to H_4_.

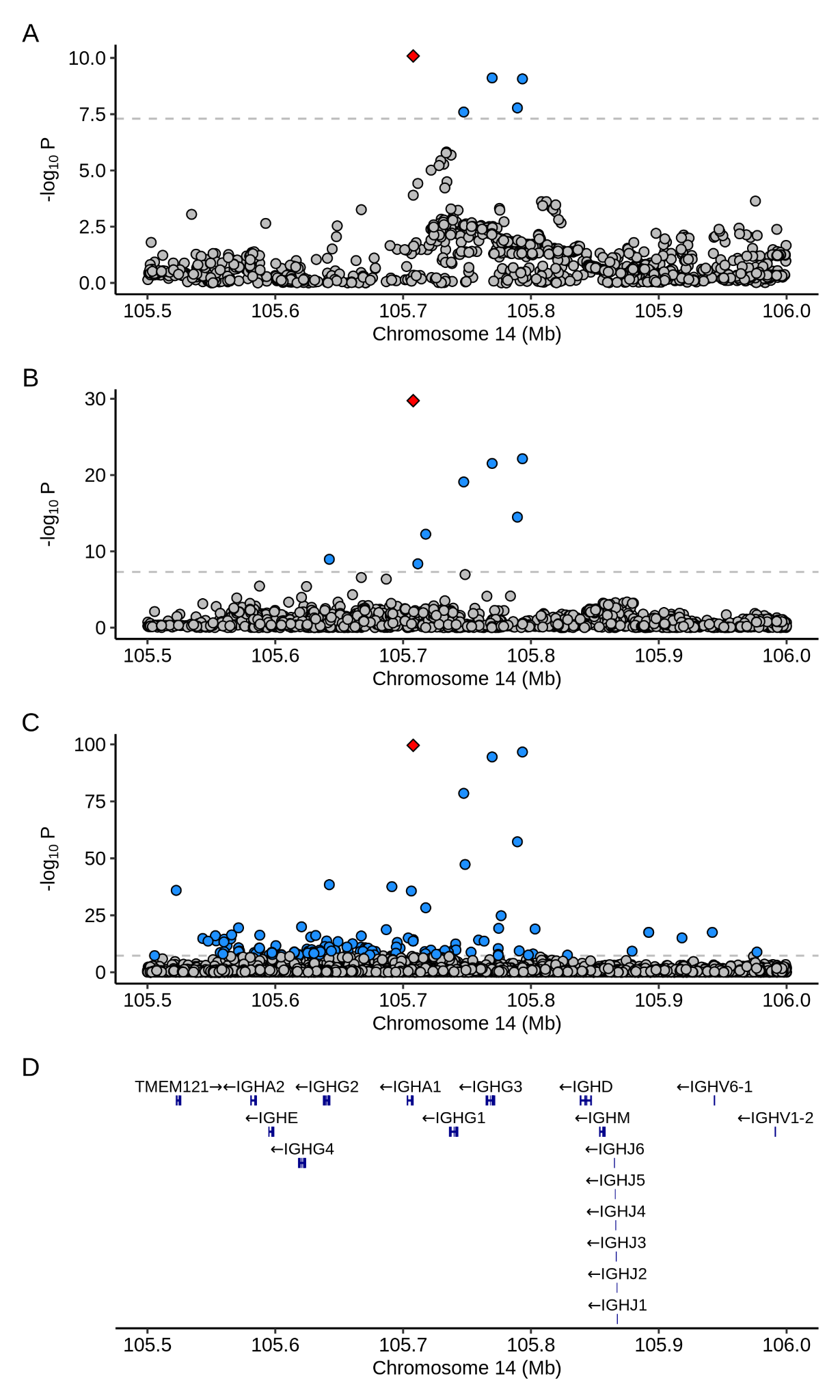

**Supplementary Figure 4**. SNP p-values in the *IGH* locus from the GWAS of serum IgA in the EPIC cohort (A), and the studies by Pietzner et al. (B) and Eldjarn et al. (C). The bottom plot (D) depicts the genes at the *IGH* locus; the numerous IGH diversity gene segments are omitted for the sake of clarity.

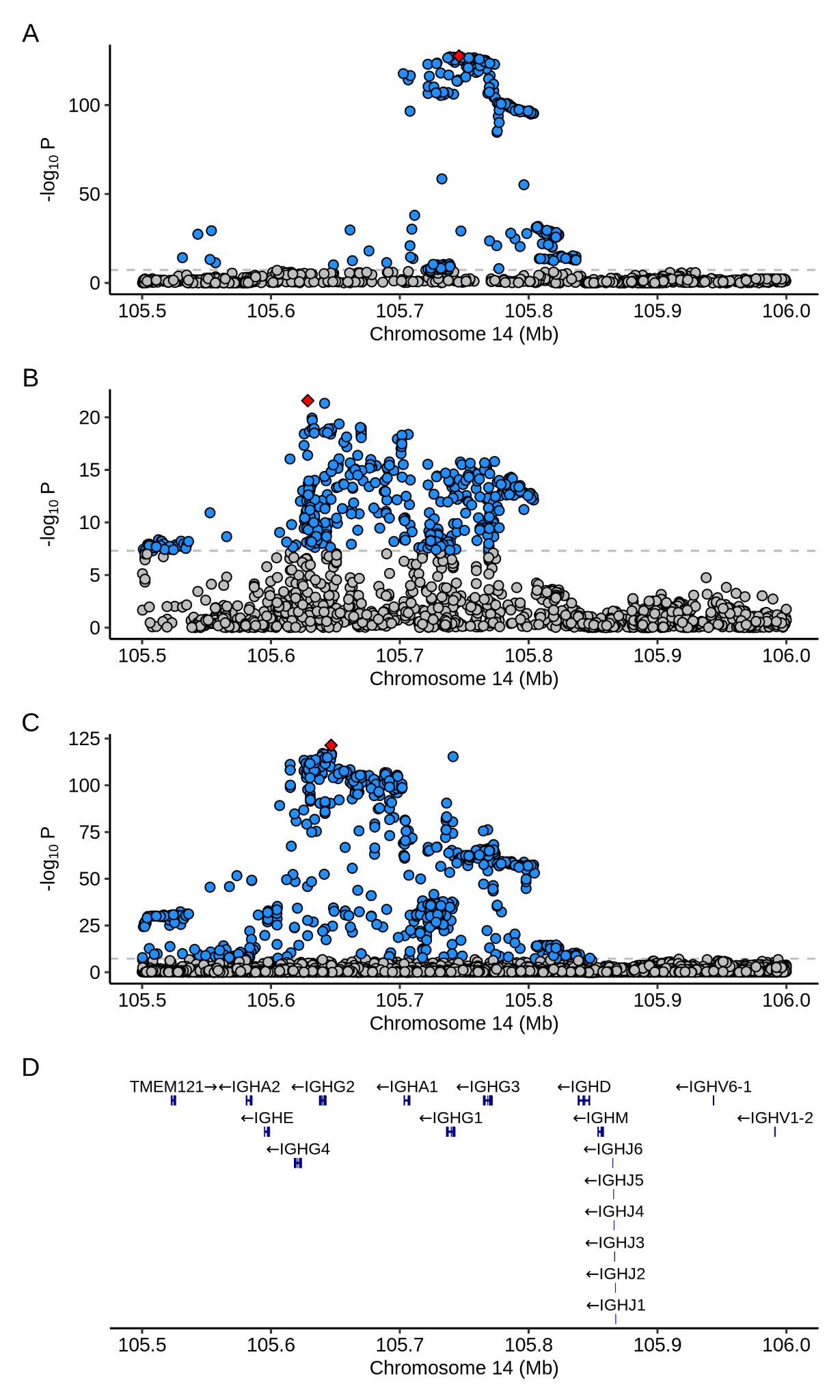

**Supplementary Figure 5**. SNP p-values in the *IGH* locus from the GWAS of serum IgG in the EPIC cohort (A), and the studies by Pietzner et al. (B) and Eldjarn et al. (C). The bottom plot (D) depicts the genes at the *IGH* locus; the numerous IGH diversity gene segments are omitted for the sake of clarity.

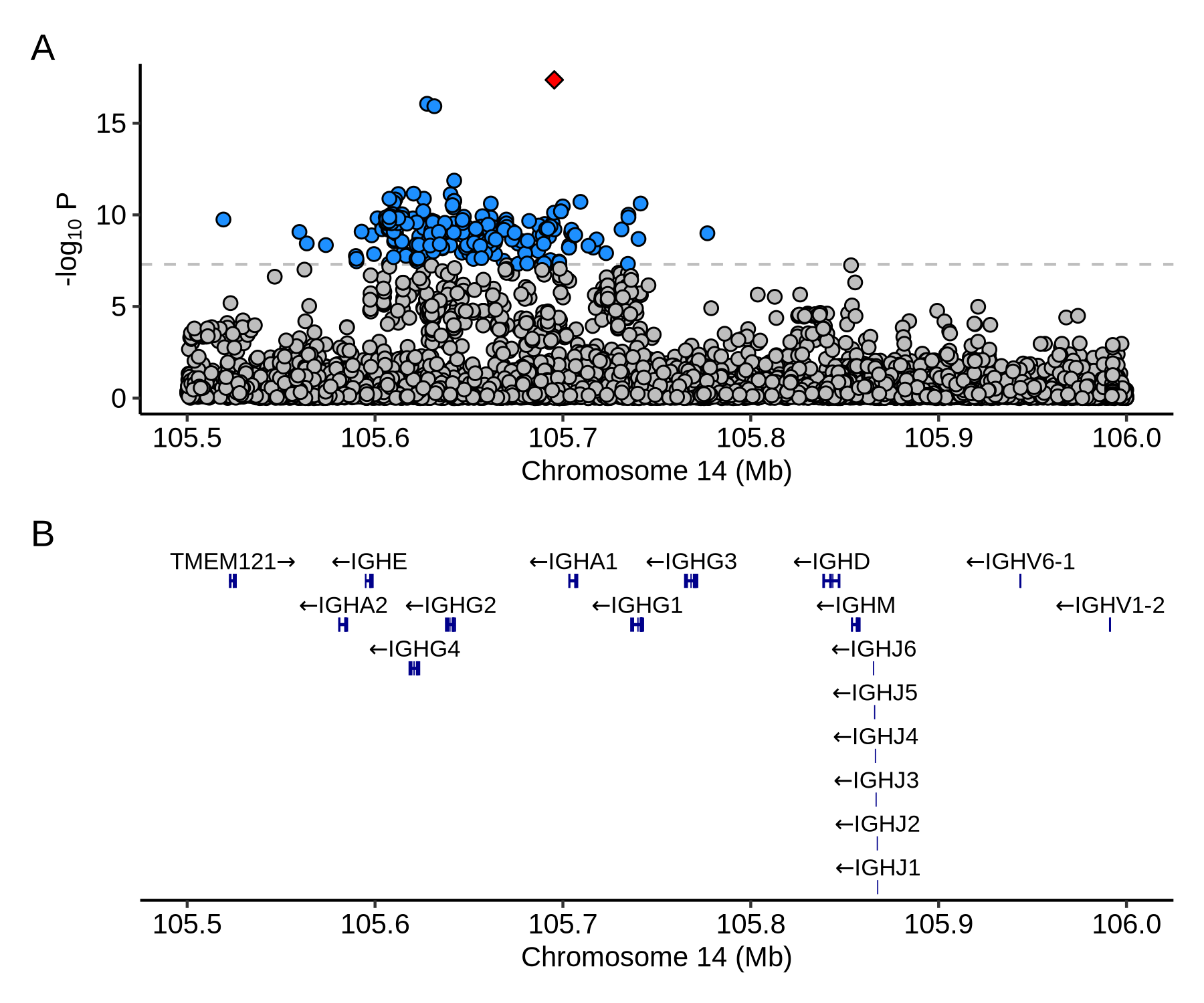

**Supplementary Figure 6**. SNP p-values in the *IGH* locus from the GWAS of serum IgM in the study by Eldjarn et al. (A). The bottom plot (B) depicts the genes at the *IGH* locus; the numerous IGH diversity gene segments are omitted for the sake of clarity.

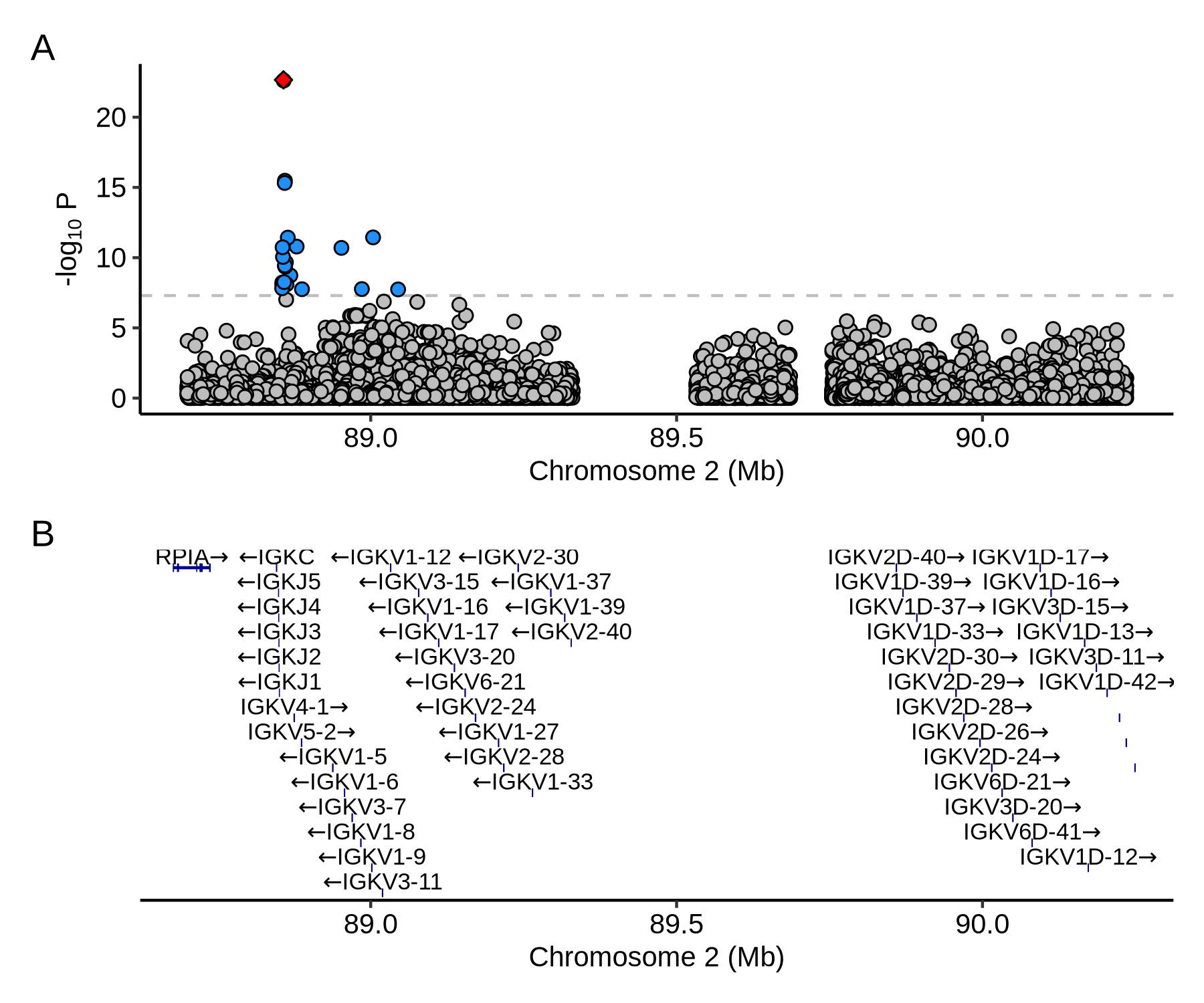

**Supplementary Figure 7**. SNP p-values in the *IGK* locus from the GWAS of serum IgG in the study by Eldjarn et al. (A). The bottom plot (B) depicts the genes at the *IGK* locus.

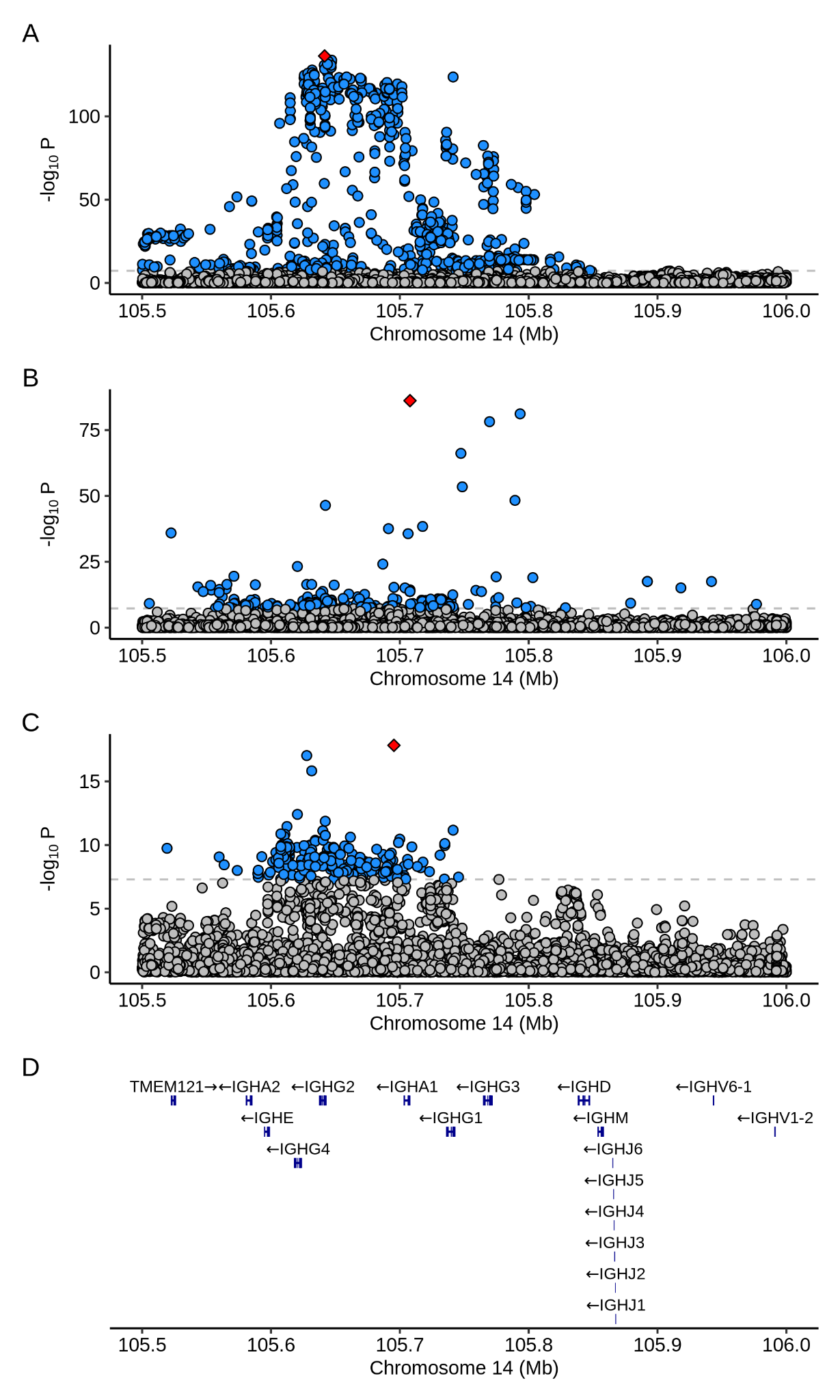

**Supplementary Figure 8**. SNP p-values in the *IGH* locus from the meta-analyses of serum IgG (A), serum IgA (B), and serum IgM (C). The bottom plot (D) depicts the genes at the *IGH* locus; the numerous IGH diversity gene segments are omitted for the sake of clarity.

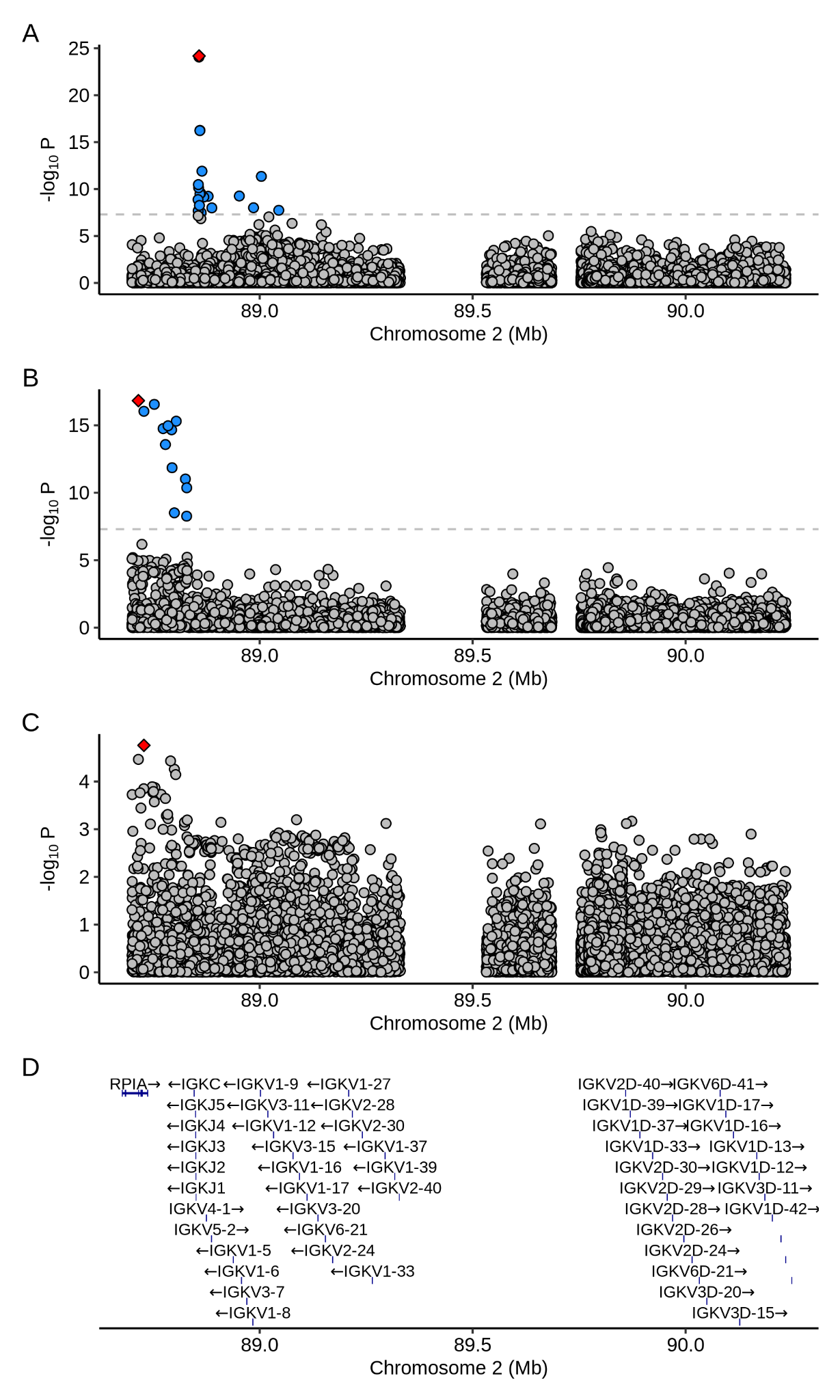

**Supplementary Figure 9**. SNP p-values in the *IGK* locus from the meta-analyses of serum IgG (A), serum IgA (B), and serum IgM (C). The bottom plot (D) depicts the genes at the *IGK* locus.

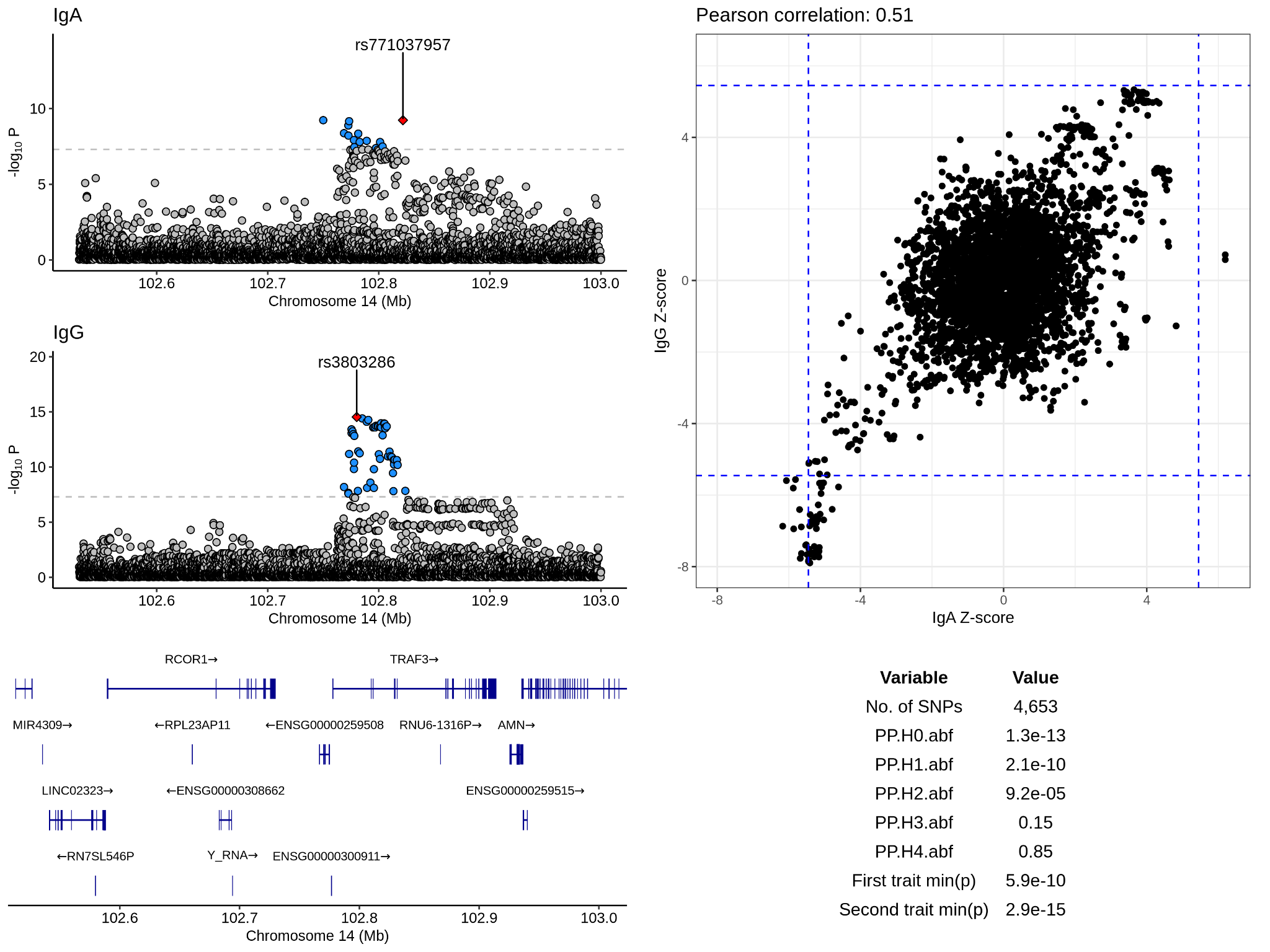

**Supplementary Figure 10.** Colocalisation analysis of the IgA and IgG association signals on chromosome 14 in the interval 102.5Mb to 103Mb. The left half of the Figure depicts meta-analytic p-values for the variants in the interval for each phenotype; lead SNPs for each phenotype’s association are coloured red and labelled. SNPs with p-values below 5x10^-8^ are coloured blue. The panel below the Manhattan plots depicts the genes located within the same genomic interval. The top panel on the right half of the Figure depicts SNP Z-scores for each phenotype. The dashed blue lines give the threshold for genome-wide significance on the Z-score scale. The table in the lower half gives statistics relating to the colocalisation analysis. ‘min(p)’ is the smallest p-value in the genomic interval subject to colocalisation analysis. The ‘PP.Hx.abf’ statistics give the posterior probability of the colocalisation hypotheses H_0_ to H_4_.

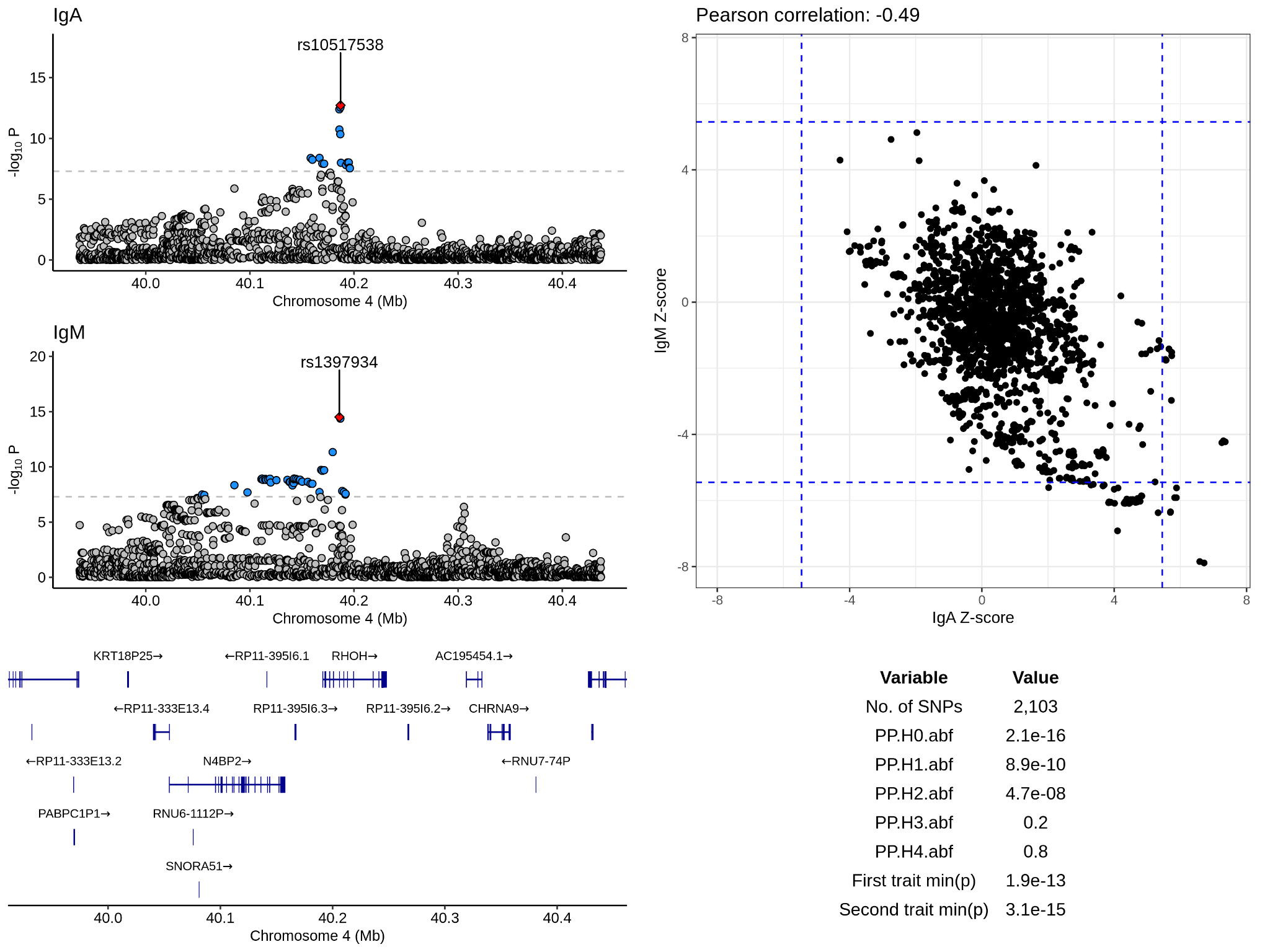

**Supplementary Figure 11**. Colocalisation analysis of the IgA and IgM association signals on chromosome 4 in the interval 39.9Mb to 40.5Mb. The left half of the Figure depicts meta-analytic p-values for the variants in the interval for each phenotype; lead SNPs for each phenotype’s association are coloured red and labelled. SNPs with p-values below 5x10^-8^ are coloured blue. The panel below the Manhattan plots depicts the genes located within the same genomic interval. The top panel on the right half of the Figure depicts SNP Z-scores for each phenotype. The dashed blue lines give the threshold for genome-wide significance on the Z-score scale. The table in the lower half gives statistics relating to the colocalisation analysis. ‘min(p)’ is the smallest p-value in the genomic interval subject to colocalisation analysis. The ‘PP.Hx.abf’ statistics give the posterior probability of the colocalisation hypotheses H_0_ to H_4_.

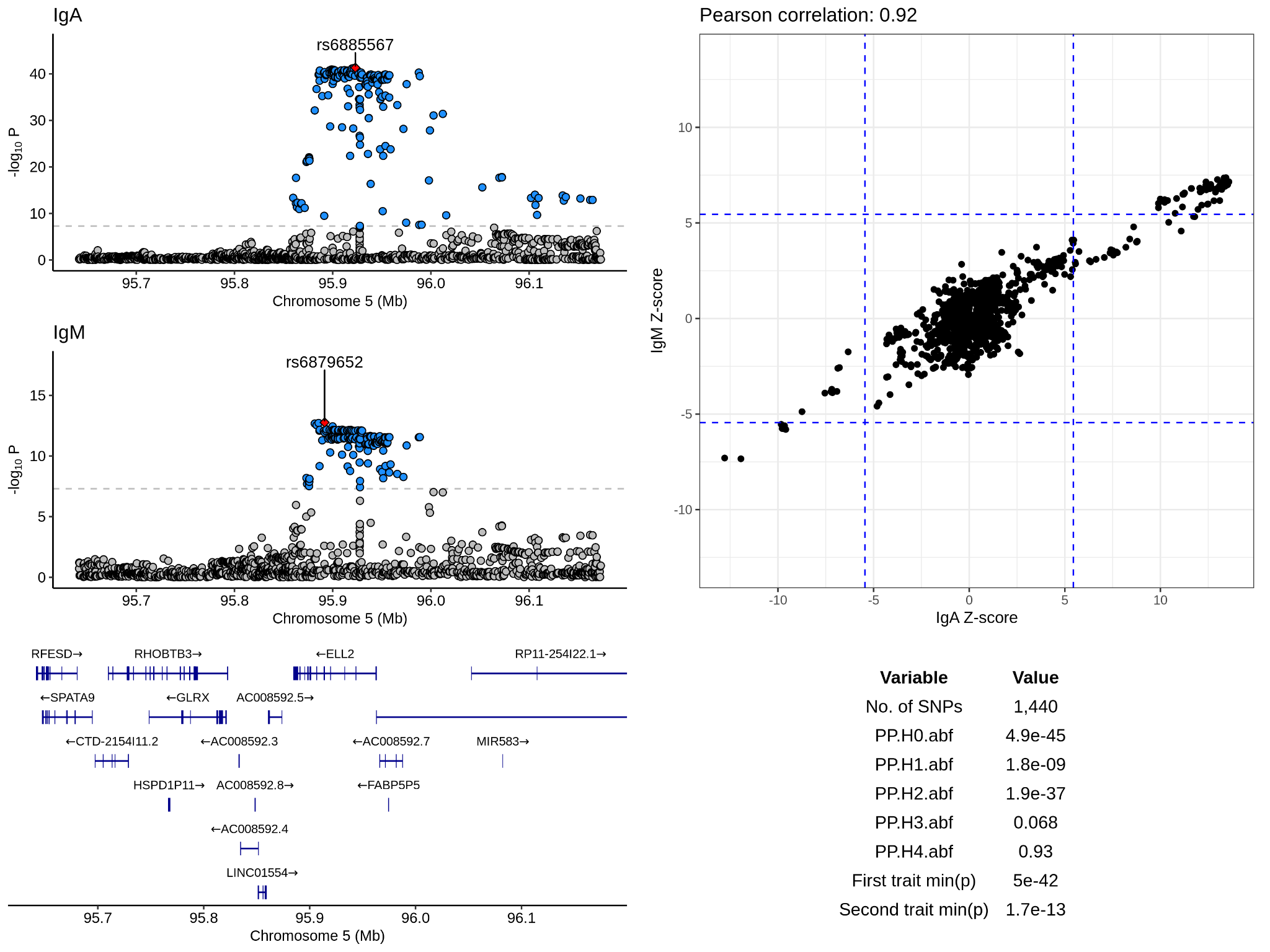

**Supplementary Figure 12**. Colocalisation analysis of the IgA and IgM association signals on chromosome 5 in the interval 95.6Mb to 96.2Mb. The left half of the Figure depicts meta-analytic p-values for the variants in the interval for each phenotype; lead SNPs for each phenotype’s association are coloured red and labelled. SNPs with p-values below 5x10^-8^ are coloured blue. The panel below the Manhattan plots depicts the genes located within the same genomic interval. The top panel on the right half of the Figure depicts SNP Z-scores for each phenotype. The dashed blue lines give the threshold for genome-wide significance on the Z-score scale. The table in the lower half gives statistics relating to the colocalisation analysis. ‘min(p)’ is the smallest p-value in the genomic interval subject to colocalisation analysis. The ‘PP.Hx.abf’ statistics give the posterior probability of the colocalisation hypotheses H_0_ to H_4_.

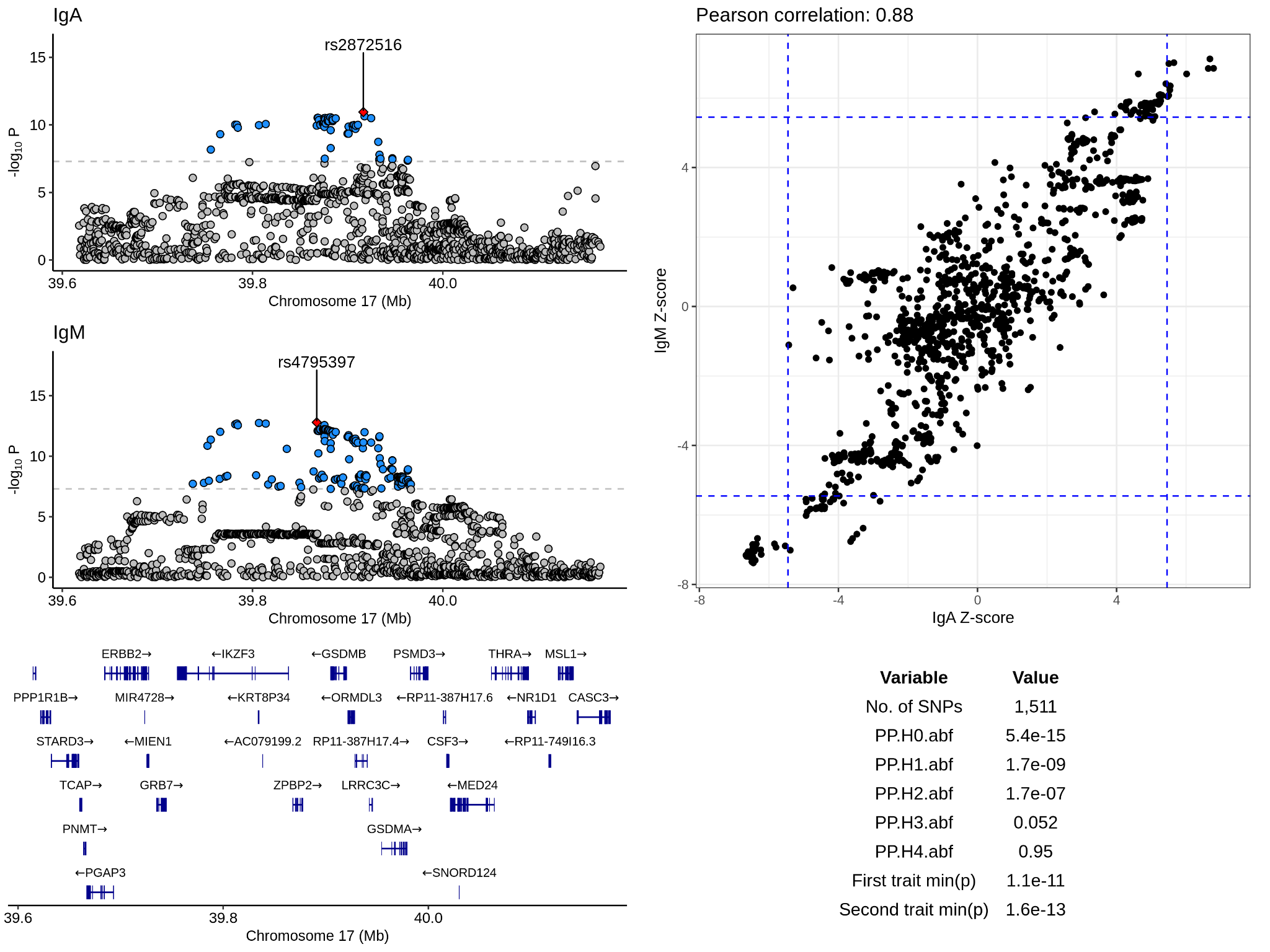

**Supplementary Figure 13.** Colocalisation analysis of the IgA and IgM association signals on chromosome 17 in the interval 39.6Mb to 40.2Mb. The left half of the Figure depicts meta-analytic p-values for the variants in the interval for each phenotype; lead SNPs for each phenotype’s association are coloured red and labelled. SNPs with p-values below 5x10^-8^ are coloured blue. The panel below the Manhattan plots depicts the genes located within the same genomic interval. The top panel on the right half of the Figure depicts SNP Z-scores for each phenotype. The dashed blue lines give the threshold for genome-wide significance on the Z-score scale. The table in the lower half gives statistics relating to the colocalisation analysis. ‘min(p)’ is the smallest p-value in the genomic interval subject to colocalisation analysis. The ‘PP.Hx.abf’ statistics give the posterior probability of the colocalisation hypotheses H_0_ to H_4_.

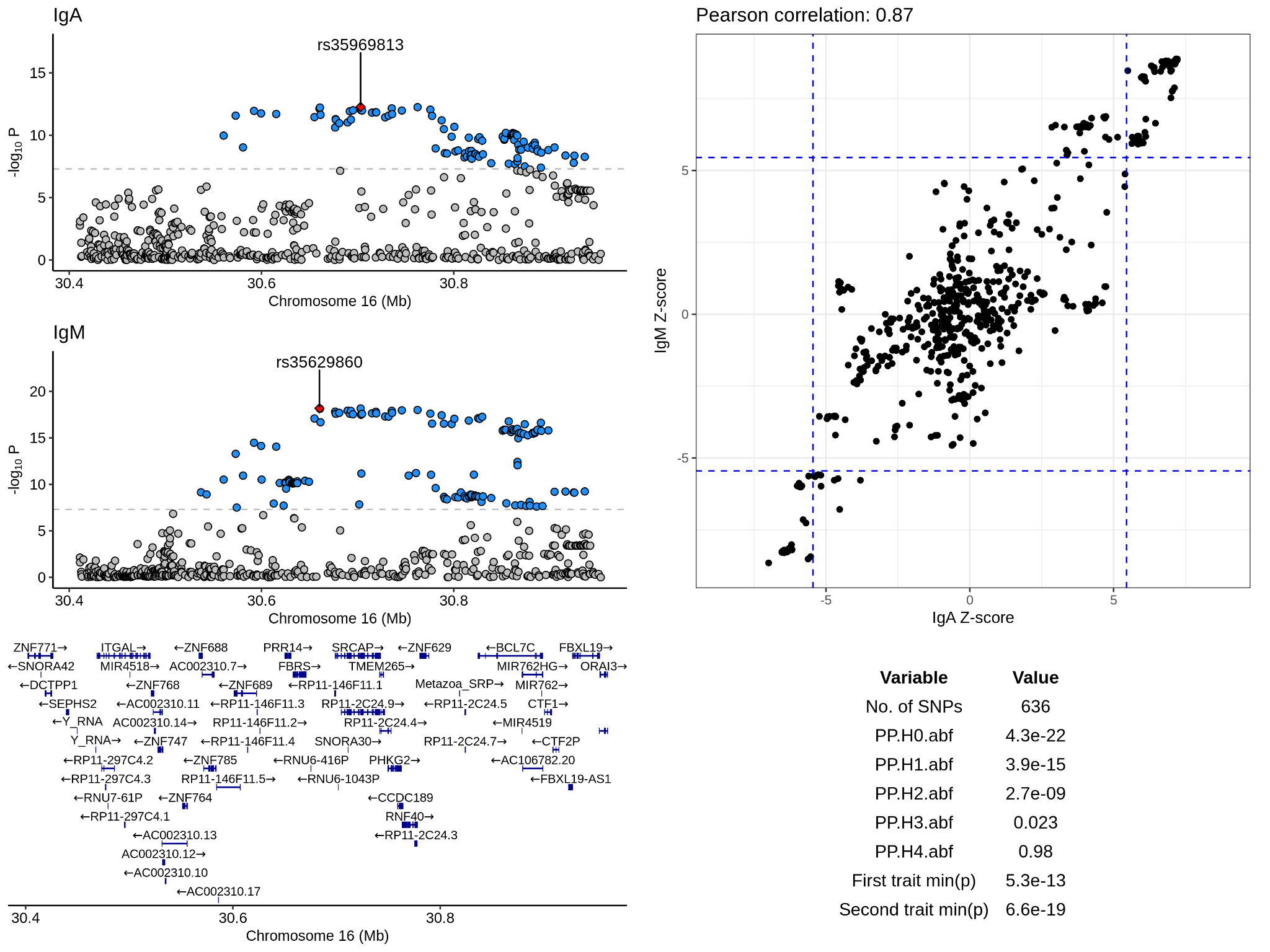

**Supplementary Figure 14.** Colocalisation analysis of the IgA and IgM association signals on chromosome 16 in the interval 30.4Mb to 31Mb. The left half of the Figure depicts meta-analytic p-values for the variants in the interval for each phenotype; lead SNPs for each phenotype’s association are coloured red and labelled. SNPs with p-values below 5x10^-8^ are coloured blue. The panel below the Manhattan plots depicts the genes located within the same genomic interval. The top panel on the right half of the Figure depicts SNP Z-scores for each phenotype. The dashed blue lines give the threshold for genome-wide significance on the Z-score scale. The table in the lower half gives statistics relating to the colocalisation analysis. ‘min(p)’ is the smallest p-value in the genomic interval subject to colocalisation analysis. The ‘PP.Hx.abf’ statistics give the posterior probability of the colocalisation hypotheses H_0_ to H_4_.

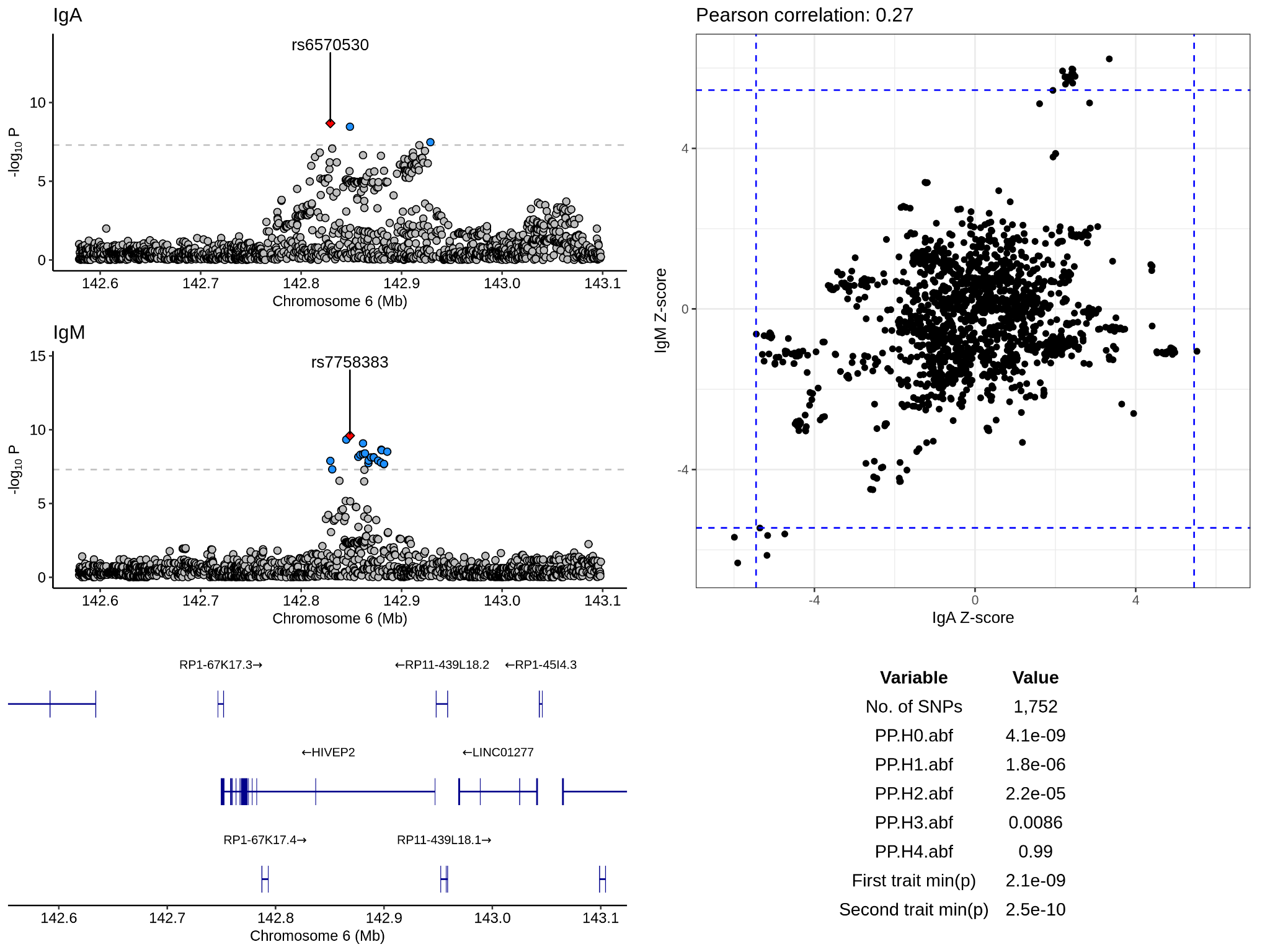

**Supplementary Figure 15.** Colocalisation analysis of the IgA and IgM association signals on chromosome 6 in the interval 142.6Mb to 143.1Mb. The left half of the Figure depicts meta-analytic p-values for the variants in the interval for each phenotype; lead SNPs for each phenotype’s association are coloured red and labelled. SNPs with p-values below 5x10^-8^ are coloured blue. The panel below the Manhattan plots depicts the genes located within the same genomic interval. The top panel on the right half of the Figure depicts SNP Z-scores for each phenotype. The dashed blue lines give the threshold for genome-wide significance on the Z-score scale. The table in the lower half gives statistics relating to the colocalisation analysis. ‘min(p)’ is the smallest p-value in the genomic interval subject to colocalisation analysis. The ‘PP.Hx.abf’ statistics give the posterior probability of the colocalisation hypotheses H_0_ to H_4_.

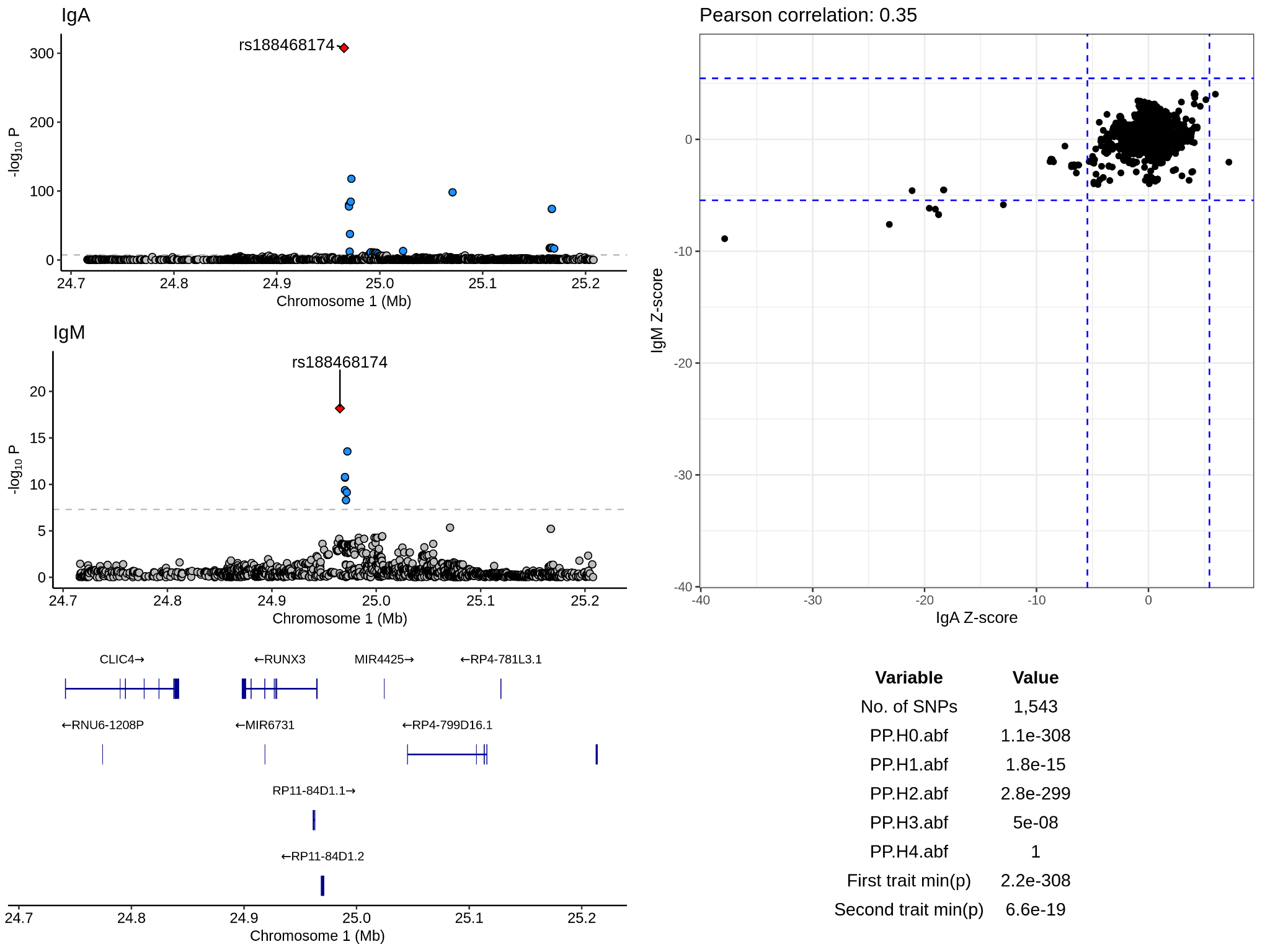

**Supplementary Figure 16.** Colocalisation analysis of the IgA and IgM association signals on chromosome 1 in the interval 24.7Mb to 25.2Mb. The left half of the Figure depicts meta-analytic p-values for the variants in the interval for each phenotype; lead SNPs for each phenotype’s association are coloured red and labelled. SNPs with p-values below 5x10^-8^ are coloured blue. The panel below the Manhattan plots depicts the genes located within the same genomic interval. The top panel on the right half of the Figure depicts SNP Z-scores for each phenotype. The dashed blue lines give the threshold for genome-wide significance on the Z-score scale. The table in the lower half gives statistics relating to the colocalisation analysis. ‘min(p)’ is the smallest p-value in the genomic interval subject to colocalisation analysis. The ‘PP.Hx.abf’ statistics give the posterior probability of the colocalisation hypotheses H_0_ to H_4_.

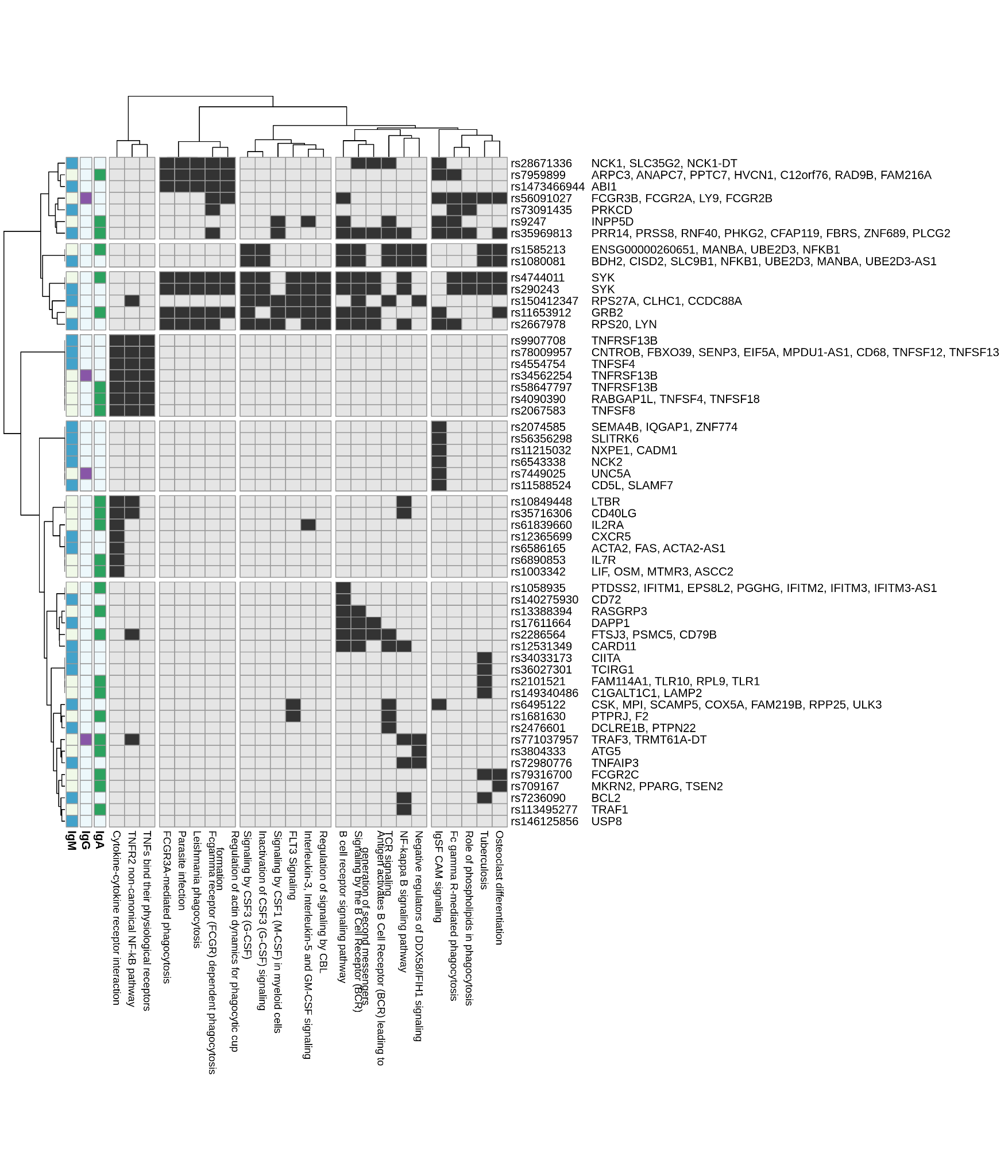

**Supplementary Figure 17.** Significant KEGG and Reactome pathways are shown together with the lead SNPs and the mapped genes which drove the association with pathway(s). Indicators on the left show which isotype(s) led to each association.

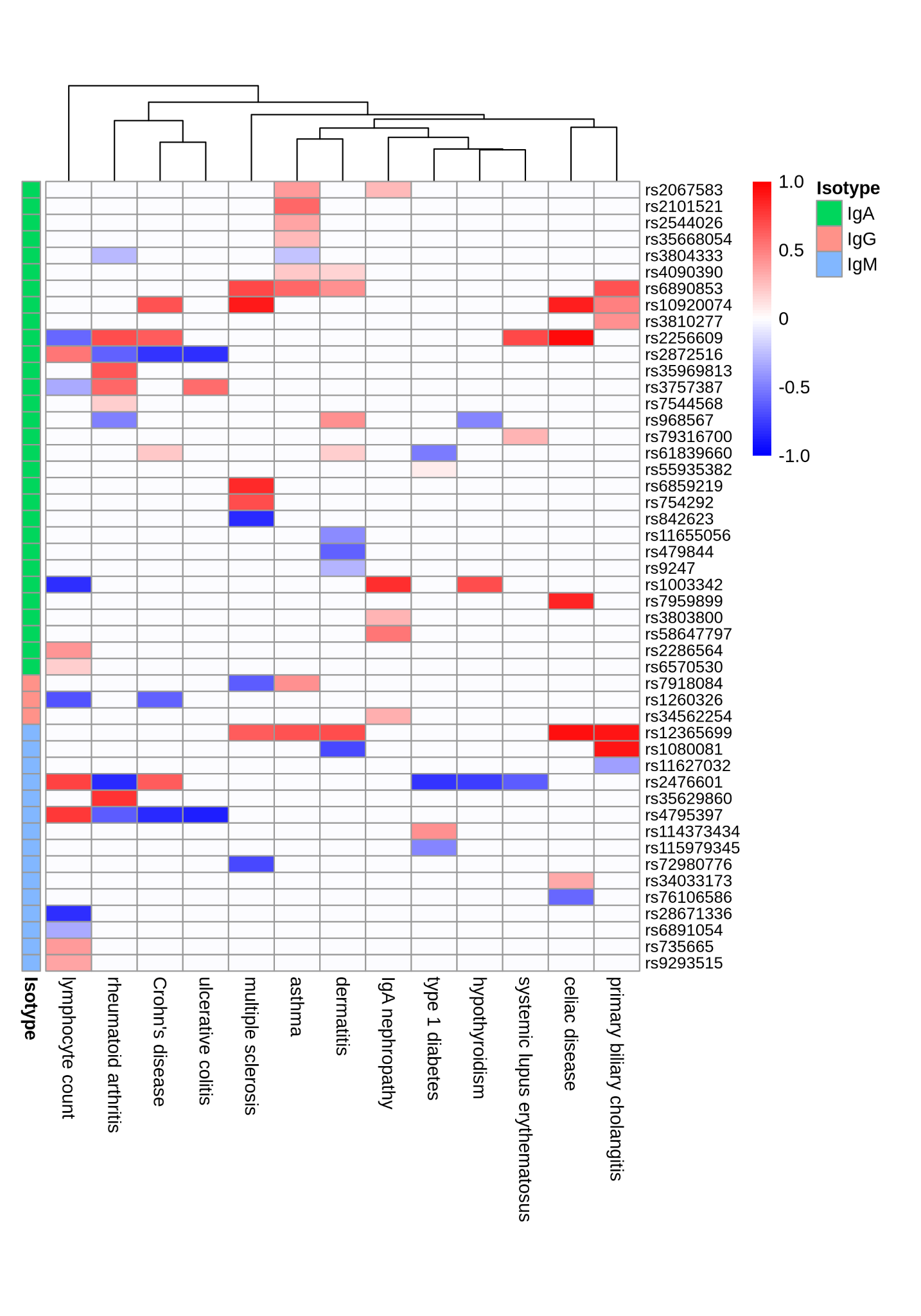

**Supplementary Figure 18**. Serum antibody associations displaying strong evidence of colocalisation with immune trait associations. The left-hand annotations give the antibody isotype. The rsIDs on the right-hand side indicate the lead SNP in the serum antibody association signal. Cell colour gives the sign of the Pearson correlation of SNP effects for the serum antibody and immune trait pair across the locus subject to colocalisation analysis. The false discovery rate, estimated as the mean of 1-PPC across colocalising loci, was 0.05.
